# Quantifying the impact of norovirus transmission in the community on acute kidney injury hospitalisations in England: a mathematical modelling study

**DOI:** 10.1101/2024.10.24.24316053

**Authors:** Hikaru Bolt, Pratik Gupte, Amy Douglas, Frank G. Sandmann, Laurie Tomlinson, Rosalind M Eggo

**Author notes:** Present address: Robert Koch Institute, Berlin, Germany (Frank G. Sandmann).

## Abstract

Norovirus is one of the most common gastrointestinal pathogens and causes substantial morbidity and mortality worldwide. A serious consequence can be acute kidney injury (AKI), characterised by a rapid deterioration of kidney function, which is associated with prolonged hospital admissions and increased mortality. This study aimed to quantify the contribution of norovirus community transmission to AKI hospitalisations in England and its attributable cost.

We developed an age-structured deterministic compartmental dynamic transmission model fitted to multiple electronic health data sources. We used a Bayesian approach to fit to hospitalisations and primary care records for AKI and gastroenteritis, as well as laboratory-confirmed norovirus surveillance data in England, from 2013-2019 and population surveys of norovirus incidence. We estimated the effect of norovirus incidence on community-acquired AKI hospitalisations and the economic burden as a product of the number of AKI hospitalisations linked to norovirus with a mean comorbidity-weighted AKI hospitalisation costs.

We estimated that 25% (95% CrI: 15-36%) of norovirus infections in people aged 65 years and above were linked to community-acquired AKI hospitalisations in the winter, amounting to 313,398 (95% CrI: 187,424-449,673) hospitalisations between 2013-2019. This represents 6.5% (95% CrI: 3.9-9.4%) of all cause community-acquired AKI hospitalisations in people aged 65 years and above over this period. The healthcare cost associated with norovirus-linked AKI admissions was estimated to be £135 million (95% CrI: £81.1 million - £195 million) annually.

Gastroenteritis has previously been identified as an important risk factor for AKI in vulnerable patients. Our results suggest norovirus incidence in the community plays an important role in driving AKI hospitalisations in individuals aged 65 years and above. This is the first population-level estimate of the prevalence of norovirus infections linked to hospitalisations for AKI. Norovirus presents a substantial economic burden on the healthcare system and AKI hospitalisation should be a consideration when evaluating future norovirus vaccines.

## Introduction

Norovirus is the most commonly reported cause of acute gastroenteritis in the UK, estimated at 3.7 million cases per year (1). While often self-limiting, with symptoms lasting 2-3 days, disease can be severe in children, the elderly, and people with impaired immune function (2). The majority of norovirus transmission is transmitted person-to-person in the community with an important burden in settings such as schools, care homes, and hospitals (3). Norovirus has a low infectious dose and is a genetically diverse virus making control efforts challenging (2,4,5). In healthcare settings like hospitals, outbreaks of norovirus cause major disruptions by prolonging admissions and due to control measures reducing bed availability during peak winter months (3,6,7). Norovirus incidence demonstrates a clear seasonality with regular peaks in winter driven by a complex interplay between population immunity, environmental factors, and host-behavioural factors (8,9).

A serious consequence of norovirus infection can be acute kidney injury (AKI). AKI is characterised by a rapid deterioration of kidney function undermining metabolic, electrolyte, and fluid homeostasis (10). Acute gastroenteritis presents with diarrhea and vomiting, leading to severe fluid losses. The resulting hypovolemia compromises renal perfusion leading to a decrease in the glomerular filtration rate: the kidneys’ ability to filter blood (11,12). It is associated with prolonged admissions and a 4-16 fold increase in the odds of death (13). In the UK, it is estimated that AKI inpatient care and longer term impacts cost £1.02 billion annually, equivalent to 1% of the NHS budget in 2011 (14). Risk factors associated with AKI include older age, chronic kidney disease, hypertension, diabetes, heart failure, drug toxicity, and is therefore typically considered primarily as a chronic disease health outcome (10). However, evidence suggests that infection is also an important contributor of AKI burden, with a substantially increased risk of AKI following gastroenteritis (12), and AKI hospital admissions demonstrating a seasonal pattern with winter increases (15–17).

With norovirus the primary cause of gastroenteritis in England, and in the absence of routine laboratory confirmation, we sought to quantify the contribution of community norovirus to AKI hospitalisations by applying mathematical modelling methods. We fit a transmission model of norovirus to AKI hospitalisation data and use Bayesian inference to estimate its contribution and attributable costs. With a number of vaccine candidates for norovirus in development (18), this study presents new insights that will serve to illustrate aspects of the potential benefits of introducing norovirus vaccination in England.

## Methods

### Transmission Model

To model norovirus transmission, we adapted a deterministic Susceptible (S), Exposed (E), Infected (I), Recovered (R) model from previously published studies (19–21). The model includes two infected compartments; symptomatic (Is) and asymptomatic (Ia) (Figure 1). The model is age stratified into four age groups: 0-4; 5-14; 15-64; 65+ years. This age stratification is to reflect young children (0-4 years) having the highest estimated incidence of norovirus, and older adults (65+ years) who are most vulnerable to severe disease (1,20,21).

**Figure 1:**
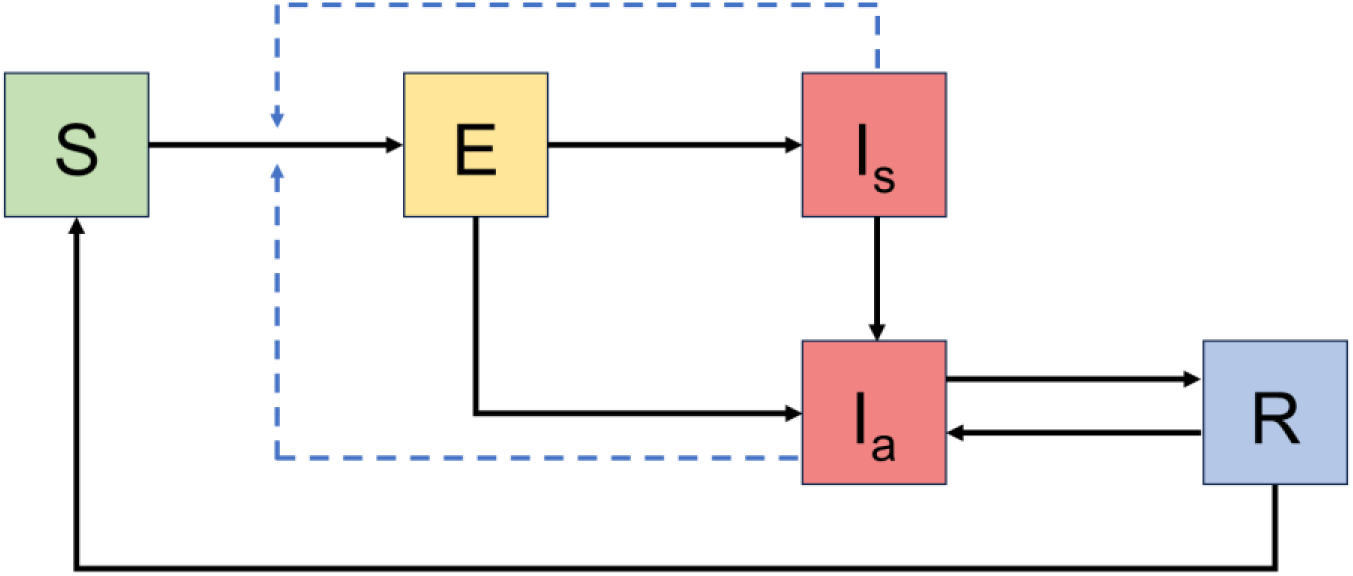
Compartmental model diagram with the following classes: *S*, susceptible; *E*, exposed; *Ia*, infectious asymptomatic; *Is*, infectious symptomatic; *R*, recovered. Arrows denote becoming infected, moving through the infection and recovery process, and waning of immunity. Force of infection is denoted by the dashed lines. Births, deaths, and ageing are included in the model but not depicted in the diagram.

Susceptible individuals are infected at rate λ into the Exposed compartment. Exposed individuals have an incubation period of μ then progress to be Infected-symptomatic (*Is*) or Infected-asymptomatic (*Ia*). To capture heterogeneous and assortative contact patterns between age-groups, denoted *cij*, we extracted contact matrix data in the UK from the POLYMOD survey using the R package socialmixr (22,23). The proportion symptomatic is σ, and the proportion asymptomatic 1-σ. Symptomatic individuals progress into the asymptomatic compartment at a rate of ψ, representing the rate of loss of symptoms. Asymptomatic individuals then progress into the Recovered compartment at a rate of γ representing the rate of recovery.

In the recovered compartment, individuals are immune and then return to the Susceptible compartment at a rate of ω. However to reflect that during the period of immunity, individuals can be re-infected but not have disease, the same force of infection applied to susceptible individuals was applied to the recovered compartment. Births, deaths, and ageing were included in the model. Individuals would age up to 80 years old, by which point they would exit the system; age-specific mortality was not applied. The full model equations, details on the force of infection and seasonality are in Supplement S1.1. Parameter definitions and values for fixed parameters are outlined in Supplement S1.3.

### Model fitting overview

We fitted our model to various sources of data which include: UK Health Security Agency’s Second Generation Surveillance System (SGSS), the second longitudinal study of infectious intestinal diseases in the UK (IID2), the Clinical Practice Research Datalink (CPRD) Aurum, and Hospital Episode Statistics (HES). Information about each data source is outlined in Supplement S2. Use of CPRD/HES was approved in 2023 (protocol 23_003034). We defined each epidemic year as running from the start of July to the end of the following June.

We assumed that each source represents distinct segments of the surveillance pyramid and captured different norovirus dynamics across the age groups (Figure 2). For example, younger age groups are more likely to be captured in GP data ; whereas in older age groups norovirus dynamics are more likely captured in hospitalisation data. This assumption therefore underpins our fitting approach: in age group 0-4, we fit our model to medically attended gastroenteritis in primary care data. In the 65+ age group we fit to gastroenteritis and acute kidney injury hospitalisation data. In the 5-14 and 15-64 age group, norovirus dynamics are poorly captured in GP and hospitalisation data, therefore we did not fit to either sources. All age groups were fitted to the SGSS surveillance data and community incidence estimates generated by IID2 (24).

**Figure 2:**
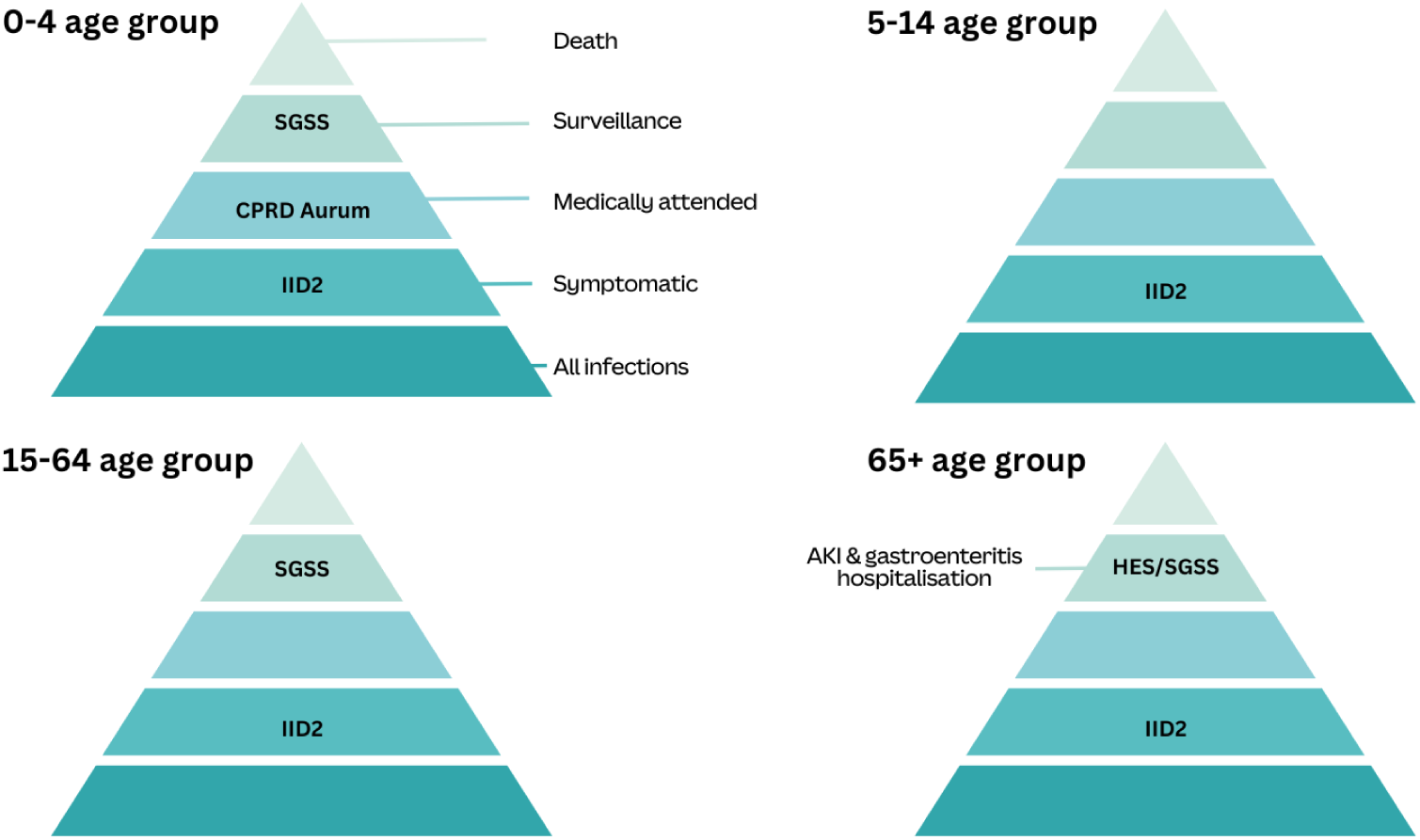
Schematic of the surveillance pyramid stratified by age. Each source of data captured different segments of the pyramid. We assumed that norovirus dynamics vary across age groups, meaning certain data sources more accurately reflect these dynamics in specific age groups. Within each age group, we label the data source used to fit the model. SGSS, Second Generation Surveillance System; CPRD Aurum, Clinical Practice Research Datalink Aurum; IID2, Longitudinal study of infectious intestinal disease in the UK (24); HES, Hospital Episode Statistics.

**Figure 2:**
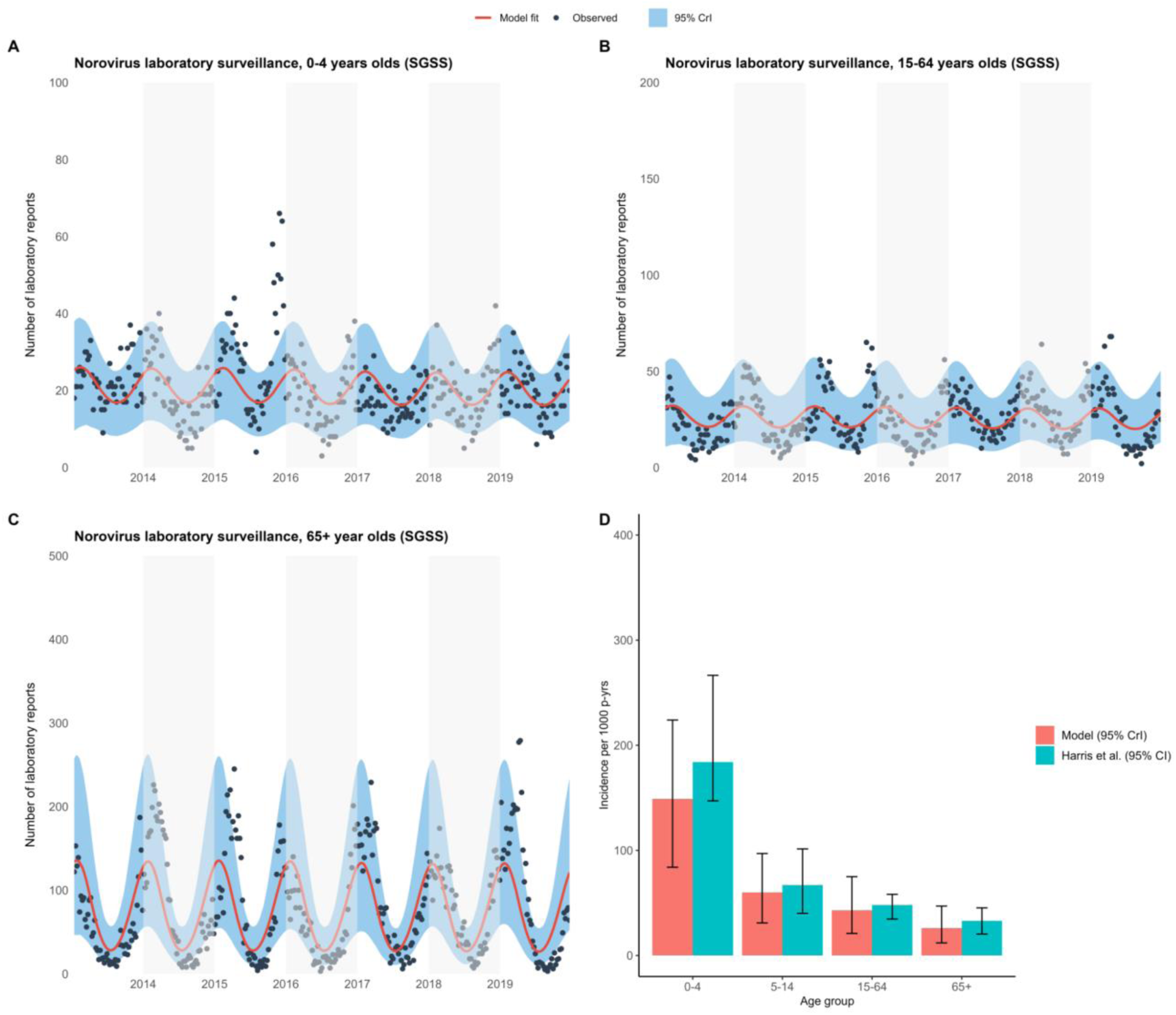
Model fit to norovirus surveillance data (SGSS), 2013-2019 stratified by age A) 0-4 years olds B) 15-64 year olds C) 65+ year olds. Black dots show the data, red the posterior median fit, and light blue the 95% credible interval. The years are shaded in grey for readability.

**Figure 3:**
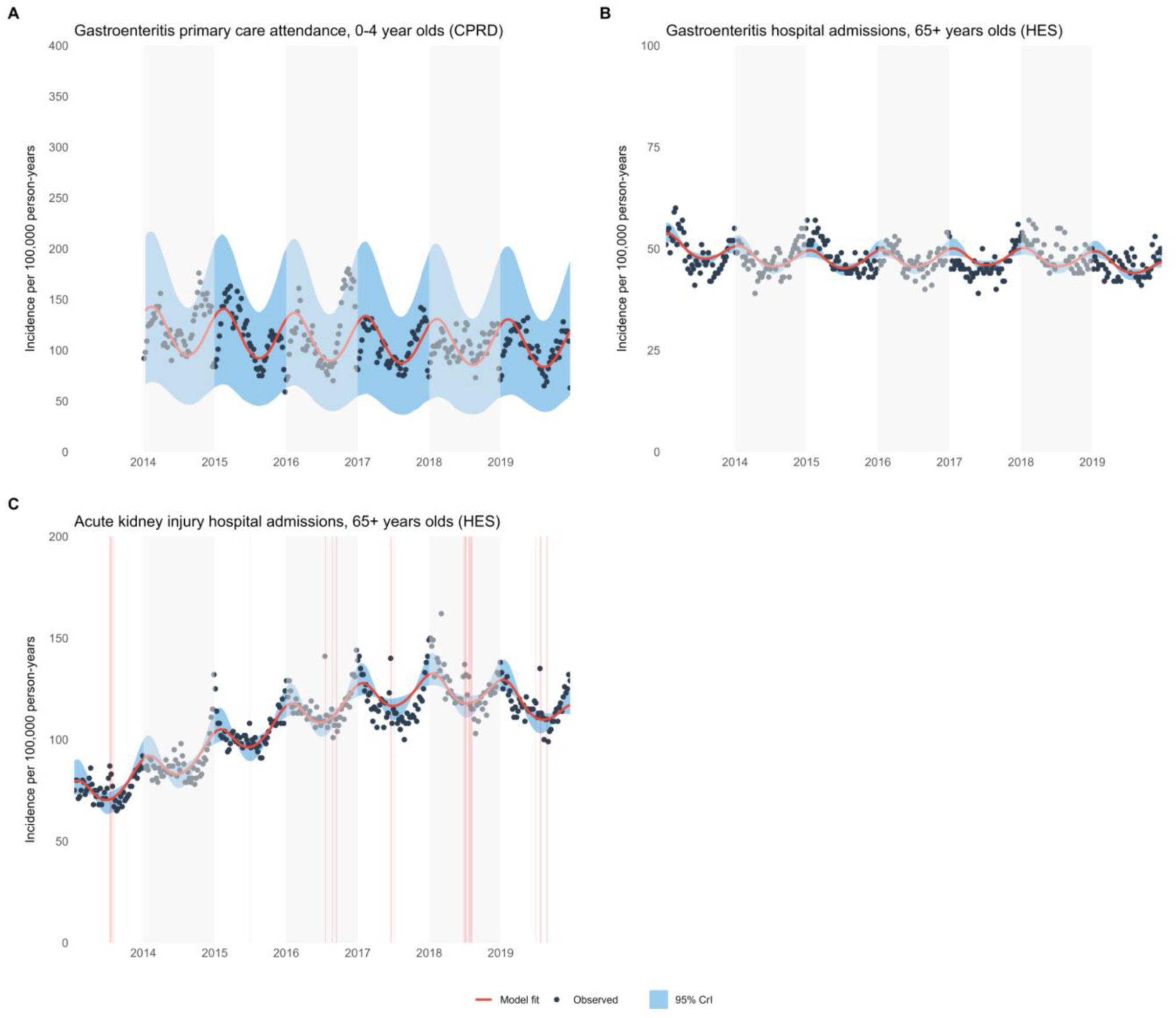
Model fit to various data sources, 2013-2019 A) gastroenteritis primary care attendance, 0-4 year olds (CPRD) B) gastroenteritis hospital admission, 65+ year olds (HES) C) acute kidney injury hospital admission, 65+ year olds (HES). Red shaded area in panel D denotes AKI hospitalisations not included in likelihood calculation as periods of AKI related to heat waves. Grey shaded area represent alternating calendar years to aid in visualising peak timings. In the 0-4 year old gastroenteritis primary care data (Panel A), we excluded data from the year 2013, as rotavirus vaccination had been introduced in the summer of 2013. Therefore to improve the specificity of the gastroenteritis attendances for norovirus, years where rotavirus were significantly more active were excluded.

We defined a GP attendance for gastroenteritis where there was a recorded pathogen code (positive test for norovirus) or a diagnosis code (e.g. gastroenteritis) (Supplement S7.3).

Symptom codes (e.g. diarrhoea and vomiting only) were not included as they were non-specific.

We defined community-acquired hospitalisations as an AKI code recorded in any position (primary, secondary, tertiary, etc.) in an episode within two days of admission, and refer to this definition as ‘AKI hospitalisation’ subsequently (Supplement S7.1). We defined gastroenteritis hospitalisation as a relevant ICD-10 code in any position in HES, at any time during an admission (Supplement S7.1).

We fitted the model to data between 2013-2019, to target a period after the introduction of the rotavirus vaccine to minimise the contribution of rotavirus as a cause of gastroenteritis. Similarly we end the observation period in 2019 to exclude the period of the COVID-19 pandemic, where norovirus dynamics were disrupted and SARS-CoV-2 was a major causative agent for AKI during that period.

### Observation model

#### Observation model for GP attendance in 0-4 age group

To parsimoniously capture secular trends we applied a B-spline function with three knots to the modelled incidence data. A B-spline function is appropriate for modelling long term patterns (25). Three knots parsimoniously captured the overall secular trend but avoided capturing seasonality of the data. The output is multiplied by an estimated parameter which scaled the modelled output to GP attendance data for all-cause gastroenteritis. A step-by-step explanation is outlined in Supplement S3.

#### Observation model for hospitalisations in 65+ year age group

To link norovirus infections to gastroenteritis and AKI hospitalisations our observation model had two components: (1) reporting parameters that fit the relationship between infections and hospitalisations, and (2) a cubic spline representing baseline AKI trends. We included a seasonal reporting parameter which allowed the relationship between infections and hospitalisations to vary between winter and summer, implemented using a smoothed cosine transition. This seasonal variation accounted for winter factors such as outbreaks in healthcare settings, higher burden in vulnerable patient populations, and potentially better detection and attribution of AKI or gastroenteritis to norovirus in the winter. We estimated both winter and summer parameters to quantify how many AKI or gastroenteritis hospitalisations are attributable to each norovirus case, and how this changed across a year (Supplement S3).

In the AKI hospitalisation data, there were short periods of increased hospitalisations in summer months strongly associated with heat waves, as defined by the UK Meteorological Office. We excluded AKI hospitalisations from the likelihood during heat waves to minimise spuriously fitting parameters to data that was unlikely to be associated with norovirus.

### Bayesian inference

We fitted parameters which govern infection dynamics (proportion of infections symptomatic, duration of immunity, probability of infection), seasonality (seasonal amplitude, seasonal offset), and reporting terms (attendance to hospital for acute kidney injury, gastroenteritis, primary care attendance, and surveillance). Fixed and fitted values, as well as priors are given in Supplement S1.3. We fitted the model to the different datasets and age groups using a quasi-Poisson likelihood (Supplement S3.4) using adaptive Metropolis-Hastings Markov Chain Monte Carlo (MCMC) . We ran four chains of 500,000 iterations, discarding 100,000 for burn in. We thinned the chain by 100 to reduce autocorrelation in the samples. We assessed convergence using trace plots, autocorrelation, effective sample size, and correlation between parameters.

### Cost analysis

To estimate the cost of norovirus-linked AKI hospitalisations we used reference costs for acute kidney injury (code LA07) reported by the National Health Service between 2013-2019 (26,27). Costs associated with AKI varied by whether an intervention was made and the complication and comorbidity score, therefore we used the mean activity weighted reference cost for each year. The mean reference cost was multiplied by the estimated number of AKI admissions linked to norovirus and a range of costs from the posterior distribution of the fitted parameter. We assume a variable cost of 15%, meaning the cost saved if the norovirus attributable AKI hospitalisation was prevented during this period (28). All costs were inflated to 2021 using the NHS cost inflation index (29).

### Sensitivity analysis

To assess the robustness of our results, we performed several sensitivity analyses. We first expanded our model to incorporate a broader definition of AKI hospitalisation wherein AKI diagnosis was not restricted to within 2 days of admission. We also assessed sensitivity to using a more stringent definition of hospitalisation data that only considered AKI codes in the top two diagnostic positions. Finally, we tested sensitivity to changes in the number of knots in the B-spline function.

## Results

### Descriptive analysis

Across six seasons between 2013 and 2019, incidence of AKI hospitalisation in 65+ years old was 80 per 100,000 person years at the beginning of 2013 and rose to 131 per 100,000 person years at the end of 2019 (Supplement S4). We observed distinct winter seasonal increases in AKI hospitalisation consistently peaking between week 52 and week 1. SGSS surveillance peak week varied between week 2 and 13 during the study period, except in the 2016/2017 season where a peak was observed in week 47. GP attendance in children 0-4 and 5-14 for gastroenteritis typically showed two annual peaks, with acute drops in attendance data in week 52; typically a period defined by Christmas/winter holidays (work and school closures) in England. GP attendances for gastroenteritis were nearly doubled each winter compared to summer. Gastroenteritis hospitalisation showed a much less pronounced winter seasonal peak compared to other sources of data, with peaks between week 2 and 13 of each season.

### Model fit and parameter estimates

In general, our model fits each source of data adequately (Figure 1 and Figure 2) and posterior parameter estimates are given in table 1. For the norovirus surveillance data, across the age groups the height and timing of the peaks and troughs were captured (Figure 1). Compared to IID2 incidence rate data, our model reported estimates within the bounds of the IID2 study estimates (Figure 1 panel D). For GP attendance for gastroenteritis and gastroenteritis hospitalisation, peaks and troughs were captured by the model (Figure 2). The fit to AKI hospitalisations captured troughs well, but the height and timing of the peaks were captured slightly less well (Figure 2 panel C).

**Table 1:** Parameters and posterior estimates.

| Parameter | Median (95% CrI) | Explanation |
| --- | --- | --- |
| Proportion symptomatic | 0.87 (0.77-0.9) | Proportion of individuals symptomatic |
| Duration of immunity | 8.9 (5.8-12) | Number of years individual is immune |
| Probability of infection between under 5s | 0.18 (0.15-0.21) | Probability of transmission in under 5s transmitting to under 5s |
| Probability of infection to over 5s | 0.041 (0.037-0.047) | Probability of transmission in under 5s transmitting to over 5s, and over 5s transmitting to over 5s |
| Seasonal amplitude term | 0.019 (0.016-0.021) | A term forcing the amplitude of the periodicity in the contact rate |
| Seasonal offset term | 0.21 (0.08-0.34) | A term forcing the timing of the periodicity in the contact rate |
| Norovirus associated AKI hospitalisation in 65+ in the winter | 0.25 (0.15-0.36) | Proportion of symptomatic norovirus infections linked to an AKI hospitalisation in over 65s in the winter |
| Norovirus associated hospitalisation in 65+ in the winter | 0.074 (0.054-0.097) | Proportion of symptomatic norovirus infections linked to a gastroenteritis hospitalisation in over 65s in the winter |
| Underreporting to surveillance 0-4 | 0.0019 (0.0017-0.0021) | Proportion of symptomatic norovirus in 0-4 year olds reported to surveillance |
| Underreporting to surveillance 15-64 | 0.00072 (0.00063-0.00082) | Proportion of symptomatic norovirus in 15-64 year olds reported to surveillance |
| Underreporting to surveillance 65+ in the winter | 0.018 (0.016-0.022) | Proportion of symptomatic norovirus in over 65s reported to surveillance in the winter |
| Underreporting to surveillance 65+ in the summer | 0.0055 (0.004-0.0074) | Proportion of symptomatic norovirus in over 65s reported to surveillance in the summer |
| GP reporting for all cause gastroenteritis in under 5s | 0.38 (0.35-0.42) | Reporting parameter linking symptomatic norovirus infection in under 5s to all cause gastroenteritis diagnosed in primary care |

**Table 2:** Estimated annual costs associated with norovirus linked acute kidney injury hospitalisations. Cost values inflated to 2021 using the NHS cost inflation index. (**29**)

| Year | Number of norovirus infections (thousands) | Number of AKI hospitalisations linked (thousands) | Mean activity weighted reference cost (£) | Annual total cost inflation adjusted (millions) |
| --- | --- | --- | --- | --- |
| 2013 | 317 | 43.1 (26-61.9) | 2,600 | £129 (£77-£185) |
| 2014 | 349 | 47.3 (28-67.9) | 2,690 | £144 (£86-£207) |
| 2015 | 334 | 45.5 (27-65.2) | 2,700 | £138 (£83-£198) |
| 2016 | 312 | 42.6 (25-61.1) | 2,610 | £123 (£73-£177) |
| 2017 | 331 | 45.9 (27-65.9) | 2,660 | £135 (£81-£194) |
| 2018 | 332 | 45.4 (27-65.1) | 2,830 | £139 (£84-£200) |
| 2019 | 318 | 43.6 (26-62.5) | 3,000 | £141 (£84-£202) |
| Total | 2,293 | 313.0 (187-450.0) |  | £949 (£568-£1,363) |

Diagnostic assessment of MCMC indicates convergence. Trace plots of parameters and multiple chains of the log likelihood show patterns of the parameter space being explored efficiently (Supplement S5). Generally priors were uninformative. The narrow posterior distributions of the probability of infection parameters suggests that the data provides strong evidence for the values estimated. The posteriors for both the duration of immunity and AKI hospitalisation parameters, though more focused than their prior distributions, still showed considerable uncertainty. The posterior for the probability of transmission between under five year olds was 0.18 (95% CrI: 0.15-0.20) and for the probability of transmission to (and between) over five years olds was 0.04 (95% CrI: 0.037-0.047) (Table 1). Duration of immunity was estimated to be 8.9 years (95% CrI: 5.9-11.8). We estimated that approximately 7.4% over 65 year olds with symptomatic norovirus infection in the winter were hospitalised (with norovirus), and 1.8% were reported to surveillance in the winter. When fitting the model to AKI hospitalisation data, we estimated that the percentage of symptomatic norovirus infections in the winter linked to an AKI hospitalisation was 25% (95% CrI: 15-36%).

### Estimate of norovirus leading to acute kidney injury hospitalisations

Between 2013 and 2019, we estimate that there were 3,117,707 (95% CrI: 2,356,661 - 4,174,546) infections in people aged 65 years and above. Based on the posterior estimate, 25.1% (95% CrI: 15.0-36.0%) were linked to an AKI hospitalisation at the peak of the winters. Across the observation period we estimate there to be 313,398 (95% CrI: 187,424-449,673) norovirus linked AKI hospitalisations. There were 4,874,534 AKI hospitalisations that meet the definition of community-acquired AKI. Given this, we estimate that 6.5% (95% CrI: 3.9-9.4%) of community-acquired AKI hospitalisations in people aged 65 years and above could be attributable to norovirus infection in 2013-2019.

### Cost analysis

We estimate that between 2013-2019, between £123 - £144 million per year was associated with norovirus linked AKI hospitalisations. This amounts to £949 million (CrI: £564 million - £1.36 billion) during the observation period. Assuming the 15% variable cost, preventing norovirus in people aged 65 years and above and thus AKI hospitalisation would amount to £142 million (Crl: £84.6-£204 million) savings between 2013-2019.

### Sensitivity analysis

Broadening the definition of AKI did not change the posterior parameter estimates (Supplement S5.8). Given that our definition of community-acquired AKI represents greater than 95% of AKI codes recorded, we would expect our results to be robust. Furthermore including hospitalisation counts strongly associated with heat waves also did not change the posterior parameter estimates. While the likelihood is increased, these only represent 20 weeks out of 365 weeks of data fitted to, therefore we would expect posterior parameter estimates to be unchanged. The posterior parameter estimates were not sensitive to the number of knots used in the observation model. Secular trends were less well captured where fewer knots were applied. We also found that posteriors were sensitive to when the definition of AKI hospitalisation code was restricted to the top two diagnostic positions (Supplement S5.8).

## Discussion

We developed a model of norovirus transmission in the community fitted to a range of norovirus-related data to estimate the percentage of infections associated with community-acquired AKI hospitalisation. We estimated that one in four norovirus infections in the winter can be linked to an AKI hospitalisation in over 65s; this accounts for 313,000 hospitalisations between 2013- 2019, with associated costs of £949 million. On an annual basis, this represents about 44,700 hospitalizations and £135 million in costs.

There are limited quantitative estimates of the role of norovirus infections in AKI, especially at a population scale. This is likely because gastroenterological symptoms may have resolved by the time of hospital admission and under recognition of the link between events. Further, there is limited virological confirmation and underascertainment of norovirus in hospital coding. Given this, most studies explore associations between gastroenteritis and AKI. For example, the prevalence of AKI among children in Uganda hospitalised with gastroenteritis was 28%, (30) while conversely among people of all ages admitted with AKI in Malawi 19.6% had gastroenteritis (31). A large Japanese study using routine data to investigate the seasonality of AKI hospitalisations found that gastroenteritis was the third most common diagnosis code on admission (12.8%) (16). Because our focus lies on estimating the population-level contribution of norovirus to AKI hospitalisations, these studies are limited in serving as comparators for our estimates. However, a self-controlled case series analysis of patients in the UK taking anti- hypertensive drugs found that individuals were 43 times more likely to experience an AKI admission following an episode of gastroenteritis than pre-infection, substantially greater than with pneumonia or urinary tract infections (12). Given this high risk observed in vulnerable individuals, our finding of a high proportion of norovirus-linked AKI hospitalisations among older adults is consistent in this context.

We found that norovirus plays an important role in the seasonality of community-acquired AKI hospitalisations. However, given the multifactorial nature of AKI, other mechanisms not captured by the model are likely to play a role and may contribute to the misalignment in the timing of norovirus peaks and that of AKI hospitalisations. Other infections and important AKI-associated comorbidities such as heart failure and myocardial infarction also demonstrate seasonal associations (32–34). Detection of AKI may also vary seasonally, which may be a plausible explanation of the consistent timing of the peaks of AKI admissions.

We found that up to 25% of norovirus infections in the winter are linked to an AKI hospitalisation in people over 65, which demonstrates the substantial burden this infection places on health services. Norovirus and control efforts significantly contribute to winter pressures, and prolonged admissions due to AKI further increases the burden on hospitals. A norovirus vaccine, which is in late-stage trials, offers a mechanism to reduce the burden of AKI hospitalisations, particularly in those at higher risk. Here, we demonstrate downstream costs on AKI, which could be included in in cost-effectiveness analysis of a norovirus vaccine. Previous estimates of the economic cost of norovirus on the NHS was nearly £300 million annually, including foregone admissions (6). When considering the cost of norovirus-linked AKI hospitalisations, the economic burden of norovirus on the healthcare system would increase further.

## Strengths and weaknesses

We have used several sources of robust large-scale national data and dynamic transmission modeling to demonstrate the non-linear age-stratified dynamics fundamental to norovirus epidemiology. This approach suggests infections circulating in younger age groups serve as important drivers of AKI hospitalisations in older adult populations - a complex dynamic that would be difficult to demonstrate using other conventional methods such as time series regression. We have provided an estimate of the public health burden of the role of norovirus in AKI hospitalisations, which is challenging due to limited virological confirmation of norovirus and underascertainment in hospital coding.

Estimating norovirus-associated AKI hospitalisations relies on the reliability of the norovirus transmission model. We estimated that the duration of immunity was 8.9 years, which is comparable to previous estimates (19,20). We estimated the probability of infection to be 0.18 per contact between under 5s, and 0.04 per contact with over 5s, both within the bounds of previous estimates (20,35). The rate of hospitalisation from norovirus in the over 65s was higher than previous estimates (36). We estimated that 7.4% (95% CrI: 5.4-9.7%) of gastroenteritis admissions in older adults were attributable to norovirus, while previous estimates were <1% in the elderly. Differences can be explained by differing periods of observation, outcomes, and statistical approaches. We estimated a surveillance reporting ratio higher than that reported in the IID2 study. However we estimated a ratio of samples in 65+ year olds, where one would expect more to be reported to surveillance, compared to the all age group estimate reported in IID2.

While the fitted parameters align with previously reported estimates and our model shows strong quantitative evidence of fitting the data, several key assumptions underpin the approach. Firstly, we use a seasonal forcing term, which assumes that norovirus seasonality is driven by increased contacts during specific times of the year and that seasonal peaks occur consistently each year. Although this term was estimated during the fitting process and includes uncertainty, it oversimplifies the more nuanced mechanisms that may cause interannual variability in peak timing, as observed in surveillance data. Despite many factors influencing norovirus and AKI seasonality, the seasonal forcing term is an effective approach to summarising these patterns without adding unnecessary complexity or minimising overfitting. Previous studies have identified temperature as a driver of norovirus seasonality, suggesting it could be incorporated in future model iterations to better account for year-to-year variations.

To enhance our approach and better capture transmission dynamics, we fitted the model using multiple data sources, although these sources often involve non-specific diagnoses. With hospitalisation data, we assumed a proportion was attributable to norovirus, acknowledging that our observation model may oversimplify accounting for other causes of non-specific outcomes like AKI and gastroenteritis, as well as their seasonality.

The uncertainty demonstrated by the posterior distribution for AKI hospitalisation rate suggests that the available data was insufficient to determine these parameters with greater precision.

ICD10 codes for AKI, particularly N17 codes, have evidence of having a high positive predictive value (37). However, our data suggested that 94% of codes were recorded within 2 days of admission, meeting our definition of community-acquired AKI. This proportion is notably higher than the UK Renal Registry’s estimate of 69% for community-acquired AKI, suggesting our study may have experienced greater misclassification, contributing to uncertainty in our estimates. When we restricted our analysis to AKI recorded only in the top two diagnostic positions, the proportion of norovirus linked hospitalisations reduced substantially. This indicates that the analysis was sensitive to diagnostic position. However, AKI was listed as the primary reason for admission in a small proportion of all AKI coded admissions and may not be a true representation of AKI hospitalisations affected by norovirus. This would be in keeping with AKI being a secondary event in a complex clinic picture, but an important factor in requiring hospital treatment. Though the causal link between gastroenteritis, volume depletion, and subsequent AKI is well-established, the complex and multifactorial nature of AKI aetiology makes it challenging to model effectively.

Although our model shows a strong fit to data both qualitatively and quantitatively, it does not account for strain-specific dynamics. This limitation could affect factors like the duration of immunity, the likelihood of symptomatic infection upon reinfection, and the probability of infection. However, the study period from 2013 to 2019 was marked by stability in strain dynamics, with the GII.4 Sydney strain being the dominant capsid type in England and globally (38). Strain dynamics may become more significant in the future, esppecially when considering potentially introducing vaccination.This is also highlighted by the newly increased circulation of GII.17 during the 2023-2024 season (39).

## Conclusion

We demonstrate that norovirus incidence in the community plays an important role in driving AKI hospitalisations, and presents a large economic burden on the English healthcare system. AKI hospitalisation should be an important consideration when evaluating future norovirus vaccines.

## Declarations

### Ethics approval and consent to participate

An application for scientific approval related to the use of the Clinical Practice Research Datalink (CPRD) data was approved by the Independent Scientific Advisory Committee of the Medicines and Healthcare Products Regulatory Agency in 2023 (protocol no. 23_003034).

CPRD were already approved via a National Research Ethics Committee for purely non-interventional research of this type. The work undertaken has also had ethical approval from the London School of Hygiene & Tropical Medicine Observational/Interventions Research Ethics Committee (LSHTM Ethics ref: 22670)

### Consent for publication

Not applicable.

### Availability of data and materials

The underlying data cannot be shared publicly because of the confidential nature of the data, and access is only provided to approved researchers. Access to primary care and hospitalisation data is available on request to the Clinical Practice Research Datalink (https://cprd.com/how-access-cprd-data). We developed an R package called noromod (https://github.com/pratikunterwegs/noromod) to execute the compartmental dynamic transmission model of norovirus. Model fitting, analysis code, and results are available at: https://github.com/ehr-lshtm/acute_kidney_injury_norovirus_transmission_model.

### Authors contributors

HB, FGS, LT, RME contributed to the conception, design, analysis and interpretation of the data for the work. HB, FGS, LT, RME contributed to the acquisition and analysis of the data. HB, AD contributed to data collection, data curation, and data interpretation. HB, PG contributed to the development of software, and review of code. All authors contributed to the writing, reviewing and editing of the manuscript, approved the final manuscript, and agreed to be accountable for all aspects of the work and publication.

## Acknowledgements

We would like to acknowledge the funding support from the National Institute for Health and Care Research (NIHR) Health Protection Research Unit (HPRU) in Modelling and Health Economics, a partnership between UK Health Security Agency (UKHSA), Imperial College London, and London School of Hygiene and Tropical Medicine. The views expressed in the study are those of the authors and not necessarily those of the National Health Service, NIHR, UK Department of Health or UKHSA. The views and opinions expressed herein are the authors’ own and do not necessarily state or reflect those of Robert Koch Institute (RKI). We would also like to acknowledge Lesley Larkin, Kathleen O’Reilly, Juan Vesga, Dorothea Nitsch, and Viyaasan Mahalingasivam for their invaluable advice.

## References

1. Harris JP, Iturriza-Gomara M, O’Brien SJ. Re-assessing the total burden of norovirus circulating in the United Kingdom population. Vaccine. 2017 Feb;35(6):853–5.

2. De Graaf M, Van Beek J, Koopmans MPG. Human norovirus transmission and evolution in a changing world. Nat Rev Microbiol. 2016 Jul;14(7):421–33.

3. Koopmans M. Noroviruses in healthcare settings: a challenging problem. J Hosp Infect. 2009 Dec;73(4):331–7.

4. Glass RI, Parashar UD, Estes MK. Norovirus Gastroenteritis. N Engl J Med. 2009 Oct 29;361(18):1776–85.

5. Hall AJ. Noroviruses: The Perfect Human Pathogens? J Infect Dis. 2012 Jun 1;205(11):1622–4.

6. Sandmann FG, Shallcross L, Adams N, Allen DJ, Coen PG, Jeanes A, et al. Estimating the Hospital Burden of Norovirus-Associated Gastroenteritis in England and Its Opportunity Costs for Nonadmitted Patients. Clin Infect Dis. 2018 Aug 16;67(5):693–700.

7. Lopman BA, Reacher MH, Vipond IB, Hill D, Perry C, Halladay T, et al. Epidemiology and Cost of Nosocomial Gastroenteritis, Avon, England, 2002–2003. Emerg Infect Dis. 2004 Oct;10(10):1827–34.

8. Rohayem J. Norovirus seasonality and the potential impact of climate change. Clin Microbiol Infect. 2009 Jun;15(6):524–7.

9. Lopman B, Armstrong B, Atchison C, Gray JJ. Host, Weather and Virological Factors Drive Norovirus Epidemiology: Time-Series Analysis of Laboratory Surveillance Data in England and Wales. Pybus OG, editor. PLoS ONE. 2009 Aug 24;4(8):e6671.

10. Rewa O, Bagshaw SM. Acute kidney injury—epidemiology, outcomes and economics. Nat Rev Nephrol. 2014 Apr;10(4):193–207.

11. Kellum JA, Romagnani P, Ashuntantang G, Ronco C, Zarbock A, Anders HJ. Acute kidney injury. Nat Rev Dis Primer. 2021 Jul 15;7(1):52.

12. Mansfield KE, Douglas IJ, Nitsch D, Thomas SL, Smeeth L, Tomlinson LA. Acute kidney injury and infections in patients taking antihypertensive drugs: a self-controlled case series analysis. Clin Epidemiol. 2018;10:187–202.

13. Wang HE, Muntner P, Chertow GM, Warnock DG. Acute Kidney Injury and Mortality in Hospitalized Patients. Am J Nephrol. 2012;35(4):349–55.

14. Kerr M, Bedford M, Matthews B, O’Donoghue D. The economic impact of acute kidney injury in England. Nephrol Dial Transplant. 2014 Jul 1;29(7):1362–8.

15. Bolt H, Suffel A, Matthewman J, Sandmann F, Tomlinson L, Eggo R. Seasonality of acute kidney injury phenotypes in England: an unsupervised machine learning classification study of electronic health records. BMC Nephrol. 2023 Aug 9;24(1):234.

16. Iwagami M, Moriya H, Doi K, Yasunaga H, Isshiki R, Sato I, et al. Seasonality of acute kidney injury incidence and mortality among hospitalized patients. Nephrol Dial Transplant. 2018 Aug 1;33(8):1354–62.

17. Phillips D, Young O, Holmes J, Allen LA, Roberts G, Geen J, et al. Seasonal pattern of incidence and outcome of Acute Kidney Injury: A national study of Welsh AKI electronic alerts. Int J Clin Pract. 2017;71(9):e13000.

18. Lucero Y, Vidal R, O’Ryan G M. Norovirus vaccines under development. Vaccine. 2018 Aug;36(36):5435–41.

19. Gaythorpe KAM, Trotter CL, Conlan AJK. Modelling norovirus transmission and vaccination. Vaccine. 2018 Sep;36(37):5565–71.

20. Simmons K, Gambhir M, Leon J, Lopman B. Duration of Immunity to Norovirus Gastroenteritis. Emerg Infect Dis. 2013 Aug;19(8):1260–7.

21. Steele MK, Remais JV, Gambhir M, Glasser JW, Handel A, Parashar UD, et al. Targeting pediatric versus elderly populations for norovirus vaccines: a model-based analysis of mass vaccination options. Epidemics. 2016 Dec;17:42–9.

22. Mossong J, Hens N, Jit M, Beutels P, Auranen K, Mikolajczyk R, et al. Social Contacts and Mixing Patterns Relevant to the Spread of Infectious Diseases. Riley S, editor. PLoS Med. 2008 Mar 25;5(3):e74.

23. Funk S. socialmixr: Social Mixing Matrices for Infectious Disease Modelling.; [Internet]. 2018. Available from: https://cran.r-project.org/web/packages/socialmixr/index.html

24. Tam CC, Rodrigues LC, Viviani L, Dodds JP, Evans MR, Hunter PR, et al. Longitudinal study of infectious intestinal disease in the UK (IID2 study): incidence in the community and presenting to general practice. Gut. 2012 Jan;61(1):69–77.

25. Bhaskaran K, Gasparrini A, Hajat S, Smeeth L, Armstrong B. Time series regression studies in environmental epidemiology. Int J Epidemiol. 2013 Aug;42(4):1187–95.

26. Department of Health and Social Care. NHS reference costs [Internet]. Available from: https://www.gov.uk/government/collections/nhs-reference-costs

27. NHS England and NHS Improvement. National Cost Collection for the NHS [Internet]. Available from: https://www.england.nhs.uk/costing-in-the-nhs/national-cost-collection/

28. Roberts RR. Distribution of Variable vs Fixed Costs of Hospital Care. JAMA. 1999 Feb 17;281(7):644.

29. Jones KC, Burns A. Unit Costs of Health and Social Care 2021 [Internet]. Personal Social Services Research Unit; 2021 [cited 2024 Aug 29]. Available from: https://kar.kent.ac.uk/id/eprint/92342

30. Imani PD, Odiit A, Hingorani SR, Weiss NS, Eddy AA. Acute kidney injury and its association with in-hospital mortality among children with acute infections. Pediatr Nephrol. 2013 Nov;28(11):2199–206.

31. Evans R, Hemmilä U, Craik A, Mtekateka M, Hamilton F, Kawale Z, et al. TO021PREVALENCE, AETIOLOGY AND OUTCOME FROM COMMUNITY ACQUIRED ACUTE KIDNEY INJURY (AKI) IN SUB-SAHARA AFRICA: RESULTS OF THE MALAWI AKI STUDY (MAKIST). Nephrol Dial Transplant. 2016 May;31(suppl_1):i70–i70.

32. Stewart S, McIntyre K, Capewell S, McMurray JJV. Heart failure in a cold climate. J Am Coll Cardiol. 2002 Mar;39(5):760–6.

33. Yamamoto Y, Shirakabe A, Hata N, Kobayashi N, Shinada T, Tomita K, et al. Seasonal variation in patients with acute heart failure: prognostic impact of admission in the summer. Heart Vessels. 2015 Mar;30(2):193–203.

34. Arntz H. Diurnal, weekly and seasonal variation of sudden death. Population-based analysis of 24061 consecutive cases. Eur Heart J. 2000 Feb 15;21(4):315–20.

35. O’Reilly KM, Sandman F, Allen D, Jarvis CI, Gimma A, Douglas A, et al. Predicted norovirus resurgence in 2021–2022 due to the relaxation of nonpharmaceutical interventions associated with COVID-19 restrictions in England: a mathematical modeling study. BMC Med. 2021 Dec;19(1):299.

36. Verstraeten T, Jiang B, Weil JG, Lin JH. Modelling Estimates of Norovirus Disease in Patients with Chronic Medical Conditions. PLOS ONE. 2016 Jul 20;11(7):e0158822.

37. Tomlinson LA, Riding AM, Payne RA, Abel GA, Tomson CR, Wilkinson IB, et al. The accuracy of diagnostic coding for acute kidney injury in England – a single centre study. BMC Nephrol. 2013 Dec;14(1):58.

38. Public Health England. PHE National norovirus and rotavirus Report 09 April 2020 – Week 15 report (data to week 13) [Internet]. Available from: https://www.gov.uk/government/statistics/norovirus-and-rotavirus-summary-of-surveillance-2019-to-2020

39. Chhabra P, Wong S, Niendorf S, Lederer I, Vennema H, Faber M, et al. Increased circulation of GII.17 noroviruses, six European countries and the United States, 2023 to 2024. Eurosurveillance [Internet]. 2024 Sep 26 [cited 2024 Sep 30];29(39). Available from: https://www.eurosurveillance.org/content/10.2807/1560-7917.ES.2024.29.39.2400625

