## Supplemental material for "Quantifying the impact of norovirus transmission in the community on acute kidney injury hospitalisations in England: a mathematical modelling study"

### S1. Transmission model and parameters

#### S1.1 Force of infection and additional model details

The force of infection (λi) is a function of the probability of transmission qi, contact between age groups $c_{ij}$, the number of people symptomatically infected (Is) and asymptomatically infected (Ia), and a seasonal forcing term (κ). The probability of infection $q_i$ is different for those <5 years old and ≥5 years old to reflect the higher incidence observed in children <5 in epidemiological and modelling studies (1–3). Symptomatic ($Is_i$) and asymptomatic ($Ia_i$) individuals contribute to the force of infection. However, we assume that asymptomatic individuals are less infectious and this is reflected by a factor ρ; reflecting the previously modelled and outbreak data suggesting reduced infectiousness of asymptomatic individuals (2,4).

In the force of infection there is a seasonal forcing term denoted κ. Similar to previously reported studies, the term for the seasonal amplitude is denoted w1 and a seasonal offset term denoted as w2 (2,3,5). The seasonal forcing term is applied to the contact matrix $c_{ij}$.

Fixed parameters were birth and death rate, contacts patterns, incubation period, duration of symptoms, duration of asymptomatic shedding, proportion of infections symptomatic, relative infectiousness during asymptomatic period. Estimated parameters from Bayesian inference were the duration of immunity, amplitude and offset of seasonal forcing, and the probability of transmission in the under 5's and over 5's. Table 2 lists the fixed and estimated parameters.

#### S1.2 Model equations

$$\frac{dS_i}{dt} = bN + \delta R_i - \lambda_i S_i - dS_i + \tau S_i;$$

$$\frac{dE_i}{dt} = \lambda_i S_i - \sigma\varepsilon E_i - \varepsilon(1-\sigma)E_i - dE_i;$$

$$\frac{dIs_i}{dt} = \sigma\varepsilon E_i - \psi Is_i - dIs_i + \tau Is_i;$$

$$\frac{dIa_i}{dt} = (1-\sigma)\varepsilon E_i + \psi Is_i + \lambda_i R_i - \gamma Ia_i - dIa_i + \tau Ia_i;$$

$$\frac{dR_i}{dt} = \gamma Ia_i - \lambda_i R_i - \delta R_i + \tau R_i;$$

$$\lambda_i = q_i \kappa \sum_{j=1}^{4} c_{ij}(Is_i + \rho(Ia_i)); \qquad \text{force of infection}$$

$$\kappa = 1 + w1\, cos\left(\frac{2\pi t}{364} + w2\right) \qquad \text{seasonal forcing}$$

*i* denotes age groups: 0-4; 5-14; 15-64; 65+

*S,* susceptible*; E,* exposed; *Is*, infectious symptomatic; *Ia,* infectious asymptomatic; *R*, recovered, *N*, total population; transition parameters described in S1.3

#### S1.3 Parameters

| Parameter | Symbol | Value | Prior | Source |
|---|---|---|---|---|
| Duration of symptoms (days) | $1/\psi$ | 2 days | - | (2,6,7) |
| Duration of asymptomatic shedding (days) | $1/\gamma$ | 10 days | - | (2,7,8) |
| Incubation period | $1/\varepsilon$ | 1 day | - | (2,6,7) |
| Aging | $\tau$ | 1/365 | - | |
| Birth rate | $1/b$ | 11.4 per 1000 | - | (9) |
| Contact rate | $c_{ij}$ | See S1.4 | | (10) |
| Relative infectiousness during asymptomatic period | $\rho$ | 0.05 | - | (2,4) |
| Proportion of infections symptomatic | $\sigma$ | Fitted | Normal; mean = 0.75; SD = 0.075 | (5) |
| Duration of immunity (years) | $1/\delta$ | Fitted | Uniform; 0.5-13 | (2,11) |
| Seasonal amplitude term | $w1$ | Fitted | Uniform; 0-0.1 | - |
| Seasonal offset term | $w2$ | Fitted | Uniform; 0-0.5 | - |
| Probability of infection between under 5s | $q1$ | Fitted | Normal; mean = 0.21; SD = 0.115 | (2,12) |
| Probability of infection to over 5s | $q2$ | Fitted | Normal; mean = 0.005; SD = 0.032 | (2,12) |
| Underreporting to surveillance 0-4 | $\chi_{0-4}$ | Fitted | Uniform; 0-0.01 | (12,13) |
| Underreporting to surveillance 15-64 | $\chi_{15-64}$ | Fitted | Uniform; 0-0.01 | (12,13) |
| Underreporting to surveillance 65+ in the winter | $\theta_{winter,65+}$ | Fitted | Uniform; 0-0.06 | (12,13) |
| Underreporting to surveillance 65+ in the summer | $\theta_{summer,65+}$ | Fitted | Uniform; 0-0.1 | - |
| AKI hospitalisation in 65+ in the winter | $\theta_{AKI,winter,65+}$ | Fitted | Uniform; 0-0.75 | - |
| AKI hospitalisation in 65+ in the summer (week 27) | $\theta_{AKI,summer,65+}$ | Fixed | 0 | - |
| Gastroenteritis hospitalisation in 65+ in the winter | $\theta_{Gastro,winter,65+}$ | Fitted | Uniform; 0-0.5 | (14,15) |
| Gastroenteritis hospitalisation in 65+ in the summer (week 27) | $\theta_{Gastro,summer,65+}$ | Fixed | 0 | - |
| GP attendance for gastroenteritis in under 5s | $\zeta$ | Fitted | Uniform; 0-0.5 | (13,14) |

#### S1.4 Contact rates

We applied the following POLYMOD UK contact matrix

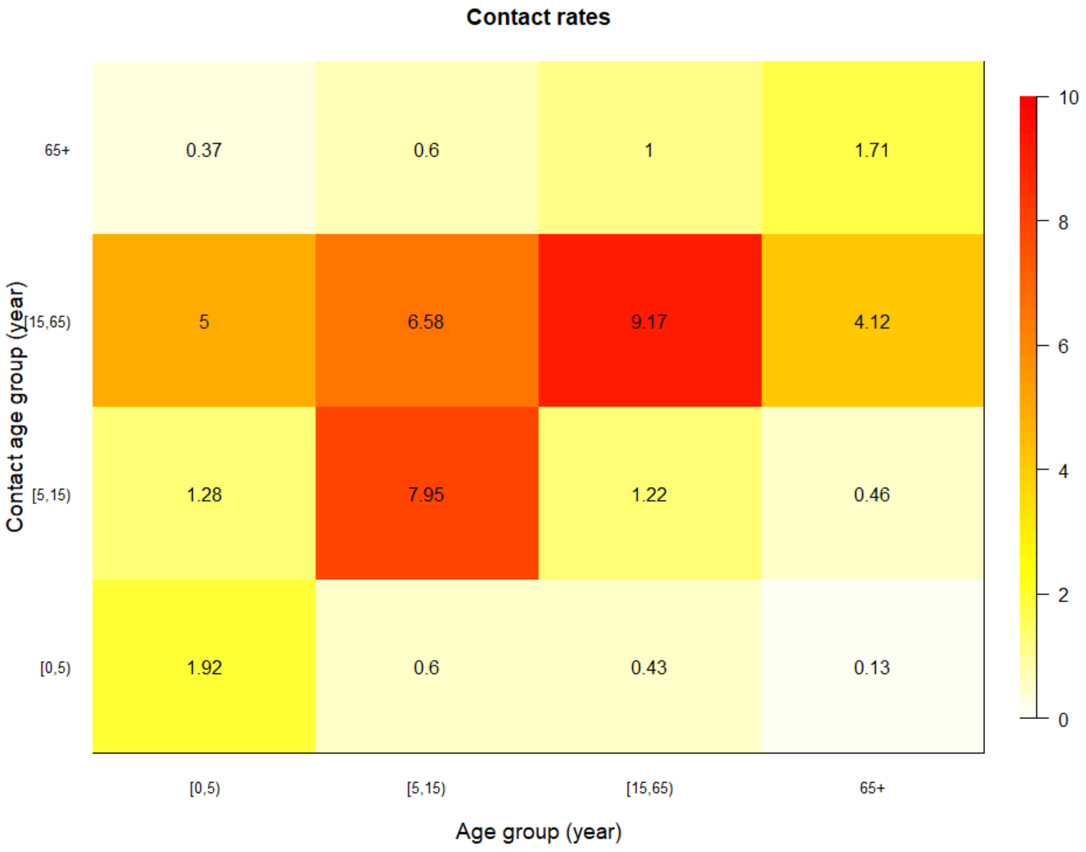

### S2. Data sources

#### S2.1 Norovirus surveillance data

We fit to data from the UK Health Security Agency's Second Generation Surveillance System (SGSS), the national laboratory reporting system for positive laboratory tests for England (16). We used deduplicated counts of laboratory data positive for norovirus from faecal and lower gastrointestinal tract specimens over time. Norovirus is not a notifiable disease, therefore surveillance in England is passive (17). Given this, samples taken represent outbreaks in care homes and health care settings, and of individuals hospitalised with more severe disease (17).

#### S2.2 Second study of infectious intestinal disease in the community (IID2)

The second longitudinal study of infectious intestinal diseases in the UK (IID2) measured incidence of sporadic community cases of laboratory positive norovirus in a prospective community cohort followed up between 2008-2009 (13). The study involved 88 primary care practices and analysed data from 6,836 participants. Compared to the general population, participants were more likely to be older, female, less deprived, and living in rural areas (13). Rates of GP consultation and reports to national surveillance were also measured. We fit our model to age stratified incidence per 1000 person years estimated by the IID2 study. We also used rates of GP consultation and under reporting to national surveillance to assess suitability of estimated under reporting parameters from the data fitting process.

#### S2.3 Clinical Practice Research Datalink (CPRD) Aurum

CPRD Aurum is a primary care database collecting longitudinal electronic health data from participating GPs representing 46 million patients with 16 million currently registered and active individuals (18). Quality assured data on patients consultations and conditions are collected using Read codes, a standardised hierarchical coding structure. Contributing general practices cover 20% of practices in the UK and the data are representative of the UK population by age and sex (19). We extracted data on attendances for gastroenteritis by age. We defined a GP attendance for gastroenteritis where there was a recorded pathogen code (positive test for norovirus) or a diagnosis code (e.g. gastroenteritis) (Supplement S7.3). Symptom codes (e.g. diarrhoea and vomiting only) were not included as they were non-specific. Attendance counts were converted into incidence per 1000 person years. We took the denominator of each year as the number of patients who were registered in the midpoint (week 27) of each year.

#### S2.4 Hospital Episode Statistics (HES)

75% of CPRD Aurum is linked to Hospital Episode Statistics (HES), which contains data on hospital admissions in England, recorded using the International Classification of Diseases version 10 (ICD-10) classification (18,19). We extracted data from HES for community-acquired AKI, and gastroenteritis hospitalisations (Supplement S7.1). We defined community-acquired hospitalisations as an AKI code recorded in any position (primary, secondary, tertiary, etc.) in an episode within three days of admission, and refer to this definition as 'AKI hospitalisation' subsequently. We defined gastroenteritis hospitalisation as a relevant ICD-10 code in any position in HES, at any time during an admission. The denominator of each year was the number of patients who were registered in the midpoint (week 27) of each year.

### S3. Model fitting

#### S3.1 Observation model explanation

*Observation model for surveillance data*

We developed an observation model to link the incidence of norovirus infections to surveillance data. For age groups 0-4 and 15-64, we simply apply a reporting parameter that scales the number of infections we simulate from the model, implying a fraction are reported to surveillance systems. This scaling factor is estimated through the Bayesian inference process.

For surveillance data reported in the age group 65+, due to factors such as outbreaks in healthcare settings, higher burden in this is vulnerable patient population, and more testing, we assume better detection to surveillance systems in the winter than the summer. We implement this by applying a reporting parameter that scales the number of infections we simulate from the model, as a smooth cosine function depicted as follows:

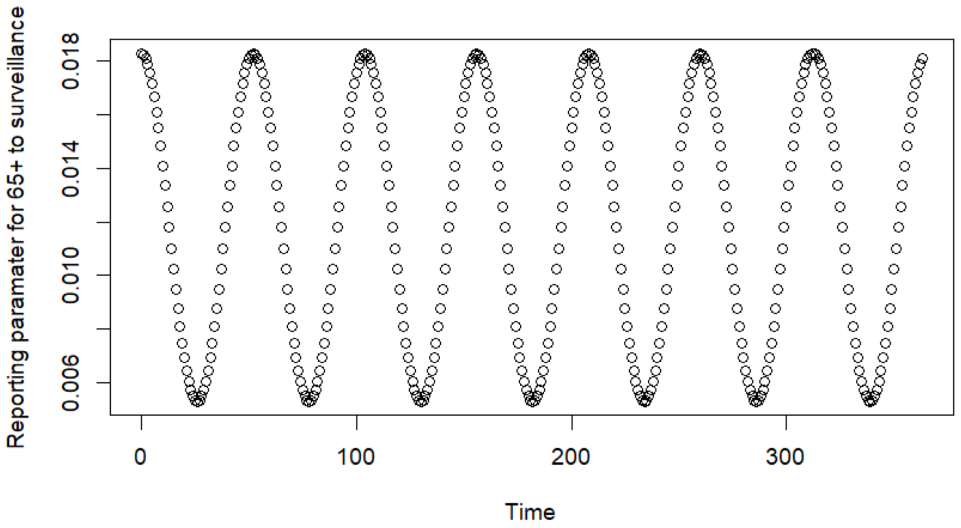

*Observation model for GP attendance*

We developed an observation model to link the incidence of norovirus infections to GP attendances for gastroenteritis in age groups 0-4. We take into account secular trends in the GP attendance data with a B-spline function of time with three knots. A B-spline function was used due to its properties of modelling long term patterns smoothly (20). Three knots were chosen, in order to parsimoniously capture the overall secular trend but avoid capturing seasonality of the data. This secular trend is then applied to the model output of norovirus infections in children aged 0-4. In the 5-14, 15-64 and 65+ age group, we assumed that norovirus dynamics are poorly captured in GP attendance data, therefore we did not fit the model to GP attendances for gastroenteritis in these groups.

The model output, now with a secular trend applied from the observed data, is multiplied by a parameter which fits the model output to observed data of GP attendance for all-cause gastroenteritis. We interpret this parameter as a factor linking a norovirus infection to all cause gastroenteritis diagnosis at a GP. This is strictly speaking not a parameter of the proportion of norovirus infections diagnosed at a GP as there are other causes of a GP attendance for gastroenteritis.

*Observation model for hospitalisations*

We developed an observation model to link the incidence of norovirus infections within the population with hospitalisations with gastroenteritis and AKI. For simplicity, we outline the process of fitting the AKI hospitalisation observation model. The method is the same for gastroenteritis hospitalisations and detailed in the visual explanation (Supplement S3.2).

Firstly, we assume that a fraction of hospitalisations are caused by norovirus infections. This is estimated through the Bayesian inference process by applying a parameter to the model output for each healthcare outcome. For example, we apply a parameter to estimate the number of norovirus infections in 65+ year olds linked to an AKI hospitalisation. However, this alone would not fit the model output to the data.

In order to link the model output to the different outcomes, we take into account additional factors influencing observations in EHR data. These factors include changes in coding practices and different disease-specific trends, all culminating in influencing non-specific disease syndromes such as AKI or gastroenteritis. To capture these trends we used a B-spline function of time with three knots on the observation data.

We then make an assumption that the sinusoidal seasonality in the AKI data is in part due to the observed seasonality in norovirus infections. In accounting for other causes of community acquired AKI, we simply add the changes in norovirus linked AKI hospitalisation incidence to the cubic spline (representing the secular trend of community acquired AKI hospitalisations). The parameter estimating the proportion of norovirus infections linked to AKI hospitalisations influences the amplitude of the model output fitting to the AKI hospitalisations data. Through Bayesian inference, we estimate the parameter values that gives the amplitude that best reflects the seasonality observed in the AKI hospitalisation data.

However, we assume that the proportion of norovirus infections linked to the AKI hospitalisation data is not a single fixed value. Due to winter factors such as outbreaks in healthcare settings, higher burden in vulnerable patient populations, we assume better detection and attribution of AKI or gastroenteritis hospitalisations to norovirus in the winter. Therefore the reporting parameter linking infection to hospitalisation data varies between the winter and summer, and the transition between seasons is captured using a smooth cosine function.

#### S3.2 Observation model visual explanation

*Observation model for GP attendance for gastroenteritis*

We took the following steps in fitting the model output to the data on gastroenteritis attendances in general practice. The figure below depicts the model output of norovirus infections in the children aged 0-4, and data on GP attendance for gastroenteritis in children aged 0-4:

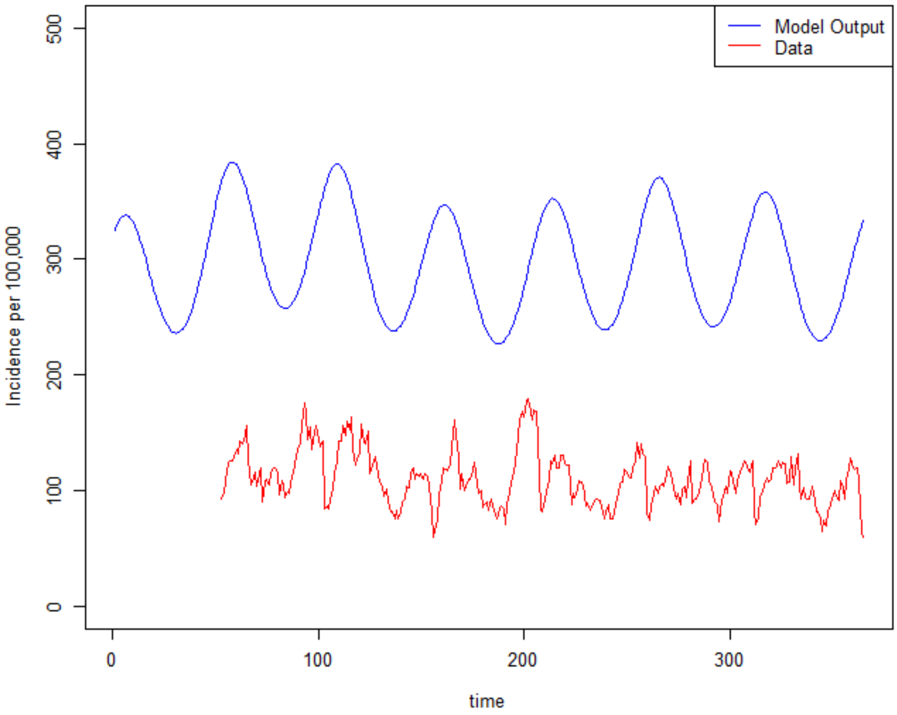

Electronic health data has long term trends which reflect changes in coding practices as well as disease specific trends which are multi-factorial when using non-specific disease outcomes such as GP attendance for gastroenteritis. In order to fit the norovirus model infections to GP attendances for gastroenteritis first we capture the long terms trends in the GP attendance data with a spline function of time. We use a B-spline basis function with 3 knots to capture the long term trend:

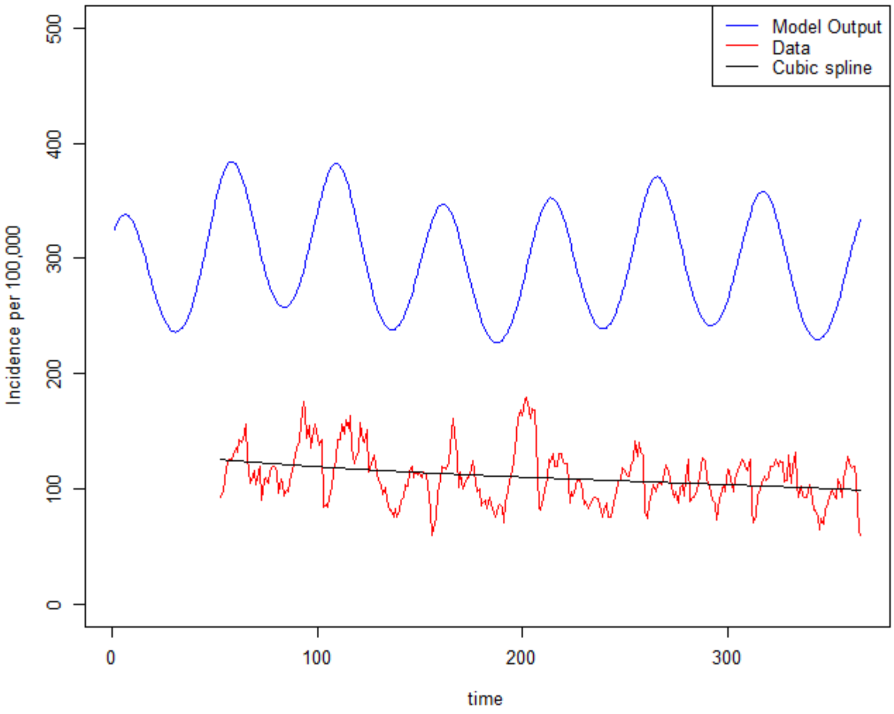

The output of the cubic spline is then added to the model output. In order to capture the changing incidence of the cubic spline over time, the median of the spline function is taken, and the difference between the spline function and the median is calculated to give and output as below:

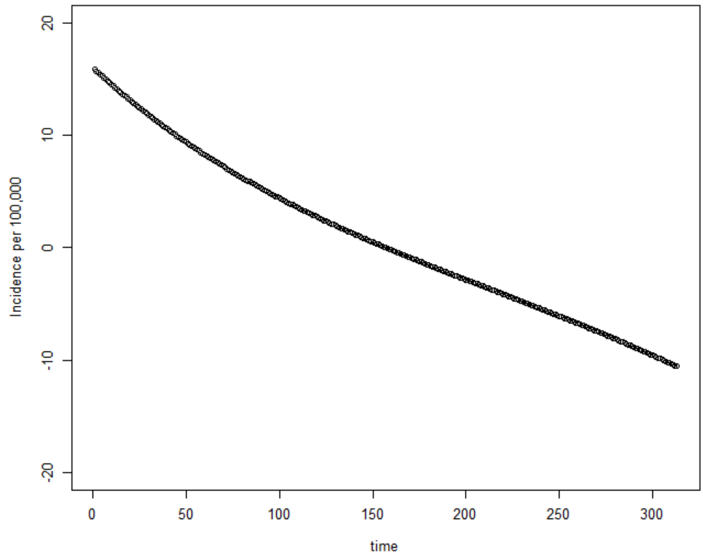

This spline function is then added to the model output to capture the long term trends observed in the GP attendance data.

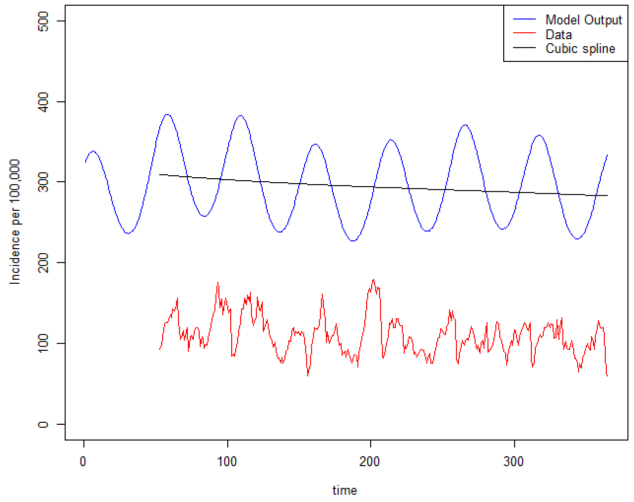

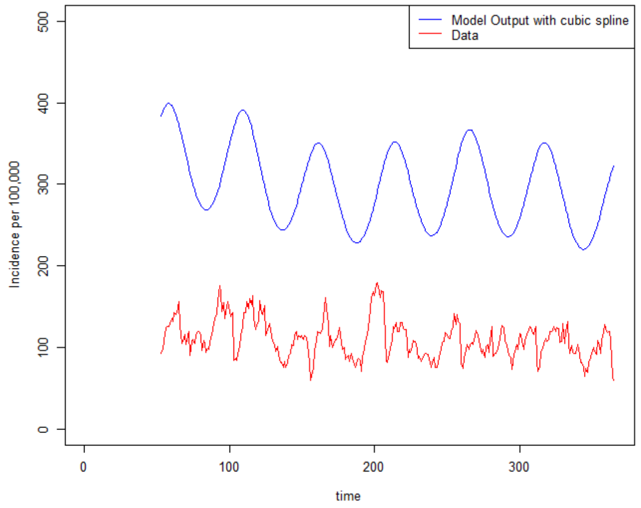

The model output with the cubic spline is then multiplied by a reporting parameter which fits the model output to observed data of GP attendance for gastroenteritis.

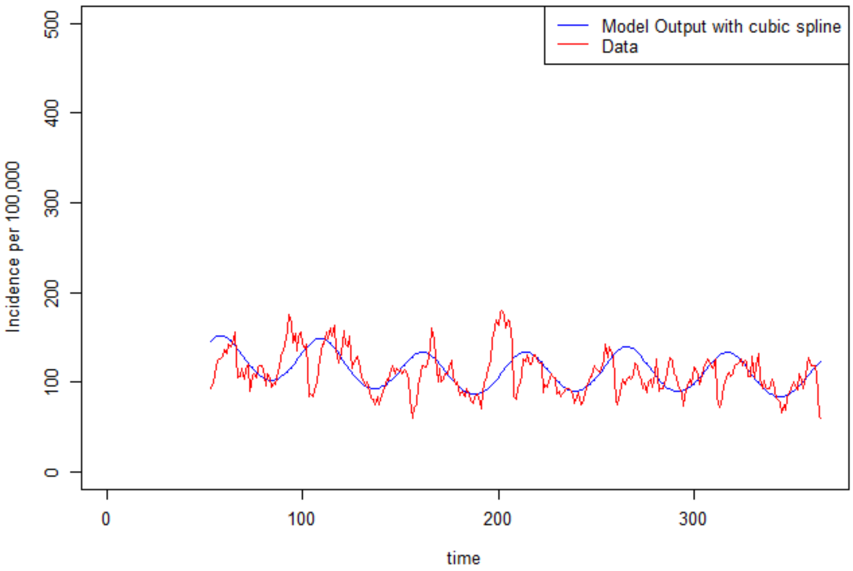

The reporting parameter is interpreted as a factor linking norovirus infections to all cause gastroenteritis diagnosis at a GP. This is strictly speaking not a parameter of the proportion of norovirus infections diagnosed at a GP as there are other causes of a GP attendance for gastroenteritis.

*Observation model for acute kidney injury hospitalisations in people aged 65 years and above*

We took the following steps in fitting the model output to the data on community acquired acute kidney injury hospitalisations.

The figure below depicts the model output of norovirus infections in the adults aged 65+, and data on acute kidney injury hospitalisations:

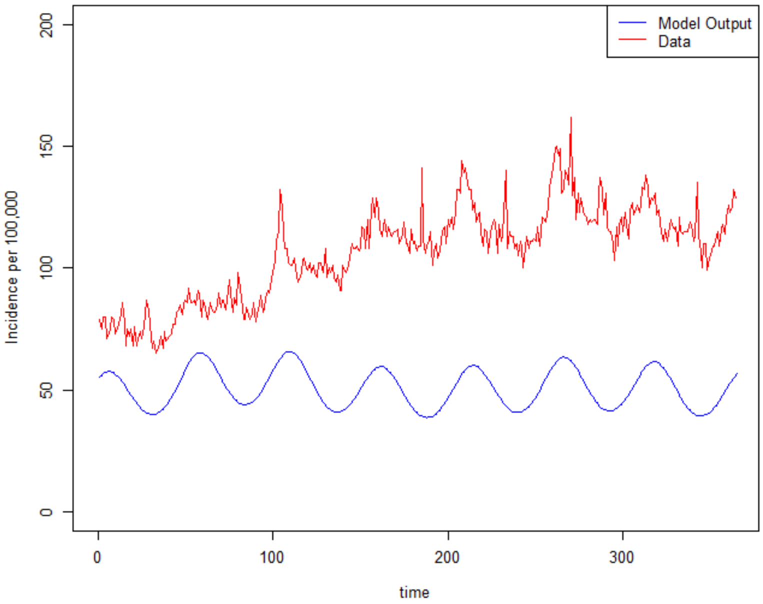

We assume that a fraction of AKI hospitalisations is caused by norovirus infections. Therefore applying a parameter, which estimates the number of norovirus infections in 65+ year olds linked to an AKI hospitalisation, we would expect the incidence of norovirus infections linked to AKI hospitalisations to be much lower than the overall incidence of community acquired AKI hospitalisations (which have many causes). We also assume that the proportion of norovirus infections linked to the AKI hospitalisation data is not a single fixed value. Due to winter factors such as outbreaks in healthcare settings, higher burden in vulnerable patient populations, we assume better detection and attribution of AKI hospitalisations to norovirus in the winter. Therefore the reporting parameter linking infection to hospitalisation data varies between the winter and summer, and the transition between seasons is captured using a smooth cosine function.

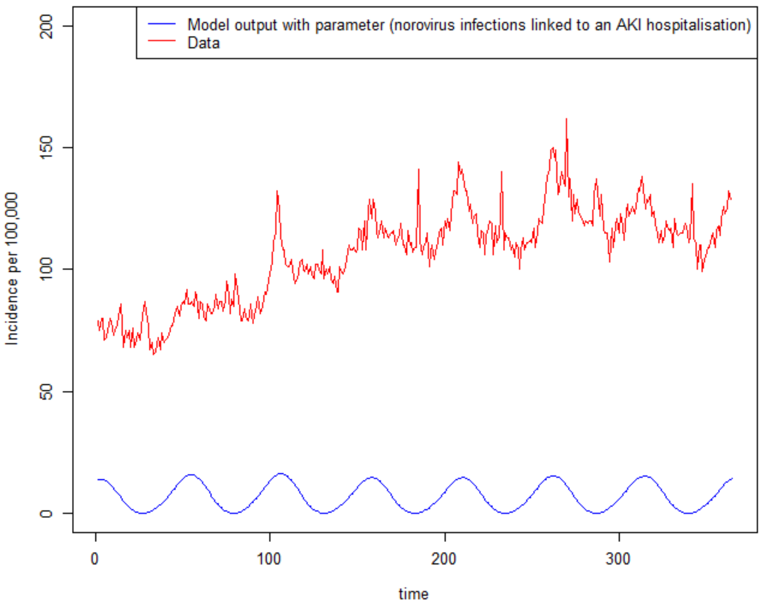

However, to fit the model output to the data on community acquired AKI hospitalisations, we need to incorporate the long term secular trend observed in the data and account for the other causes of community acquired AKI. First we capture the secular trend in the data with a B-spline basis function with 3 knots:

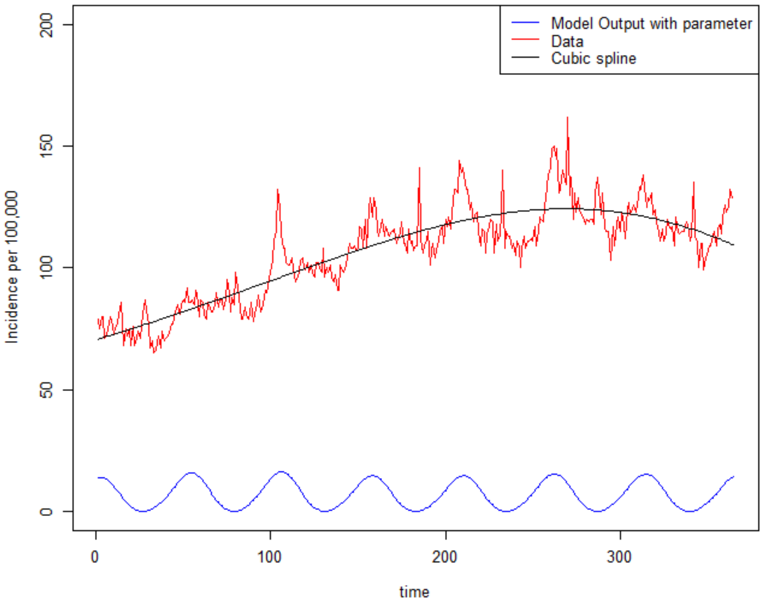

We then make an assumption that the sinusoidal seasonality in the AKI data is in part due to norovirus infections. In accounting for other causes of community acquired AKI, we simply add the changes in norovirus linked AKI hospitalisation incidence to the cubic spline (representing the secular trend of AKI hospitalisations, and community acquired AKI hospitalisations). The changes in the norovirus linked AKI hospitalisation incidence is calculated by taking the difference between the model output and the median of the model output to give an output as below.

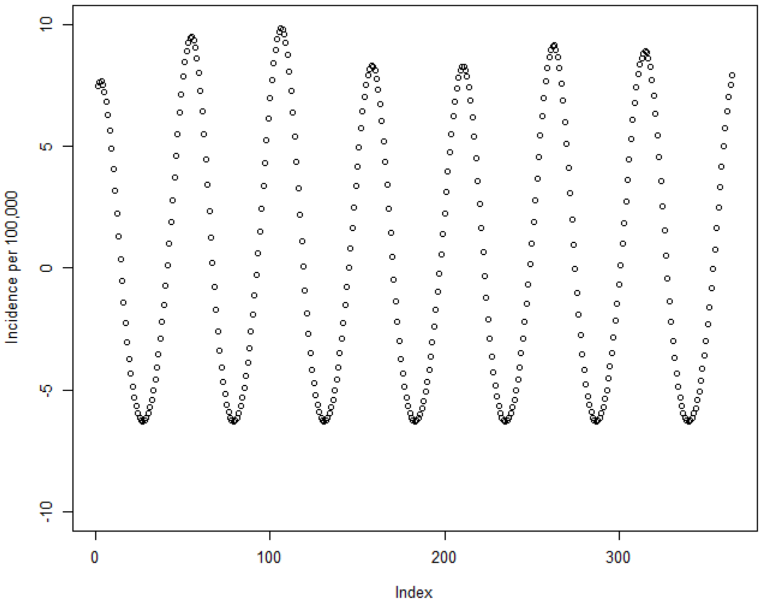

Then added to the cubic spline to fit to the data time series.

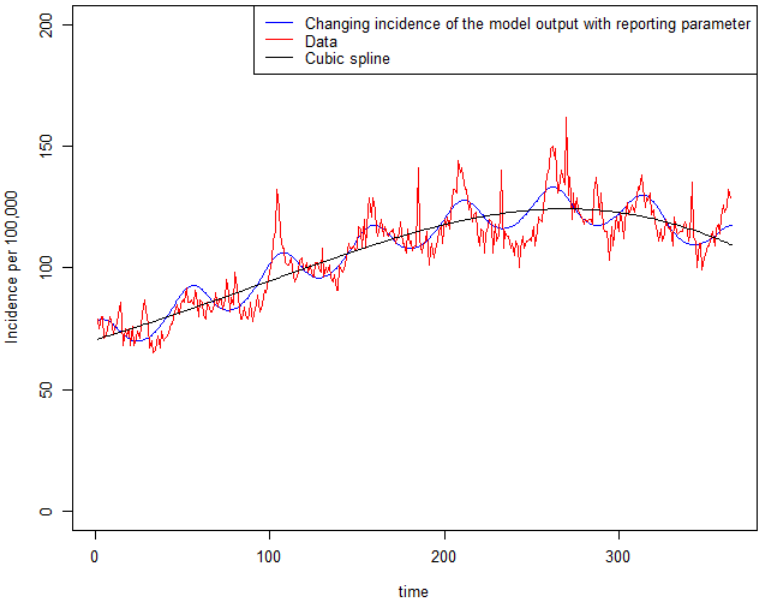

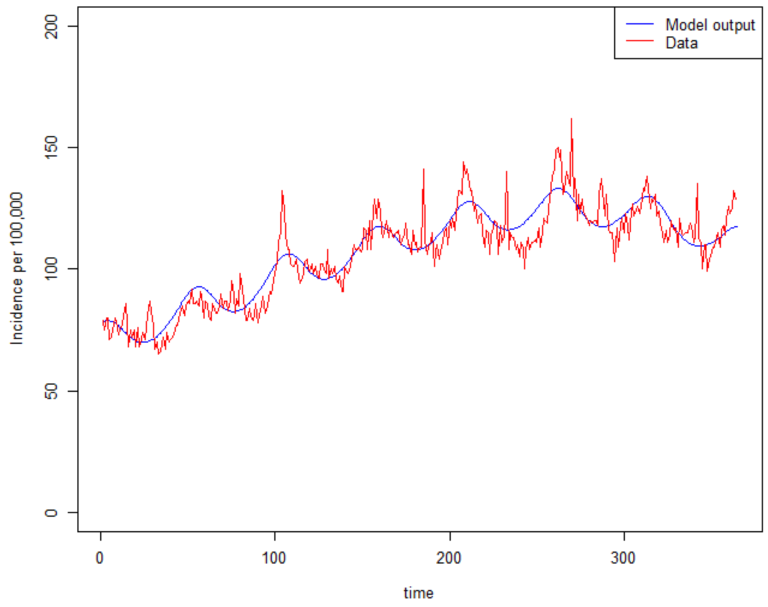

The parameter estimating the proportion of norovirus infections linked to AKI hospitalisations determines the amplitude of the model output fitting to the AKI hospitalisations data. Therefore assuming norovirus infections in part drives the sinusoidal seasonality observed in AKI, the larger proportion of norovirus linked AKI hospitalisation, then we would expect more extreme seasonality of the AKI data. Additionally, we assume that the fit between norovirus infections and all cause AKI. Through Bayesian inference, the parameter estimates the value giving the best fit to the AKI hospitalisation data.

*Observation model for gastroenteritis hospitalisations in people aged 65 years and above*

We took the following steps in fitting the model output to the data on gastroenteritis hospitalisations. The figure below depicts the model output of norovirus infections in the adults aged 65+, and data on gastroenteritis hospitalisations:

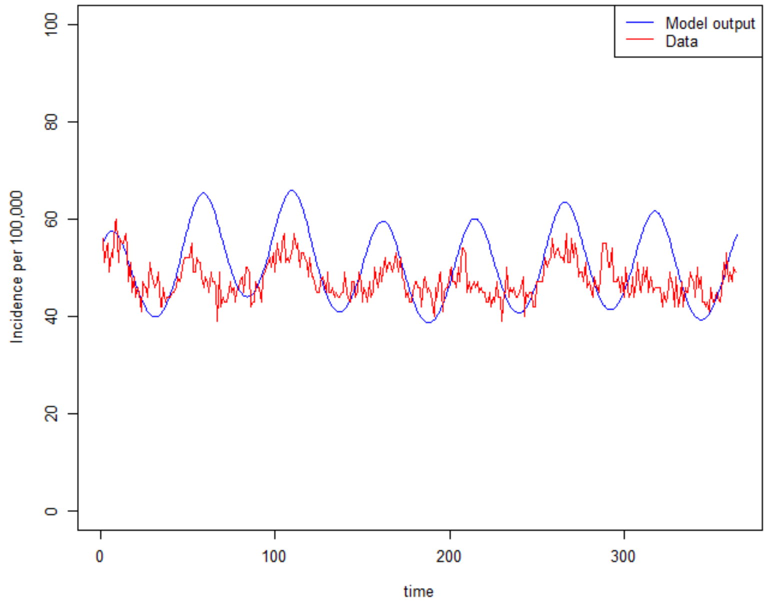

We assume that a fraction of gastroenteritis hospitalisations are caused by norovirus infections. Therefore applying a parameter, which estimates the number of norovirus infections in 65+ year olds linked to gastroenteritis hospitalisation, we would expect the incidence of norovirus infections linked to gastroenteritis hospitalisations to be much lower than the overall incidence of gastroenteritis hospitalisations (which have many causes). We also assume that the proportion of norovirus infections linked to the gastroenteritis hospitalisation data is not a single fixed value. Due to winter factors such as outbreaks in healthcare settings, higher burden in vulnerable patient populations, we assume better detection and attribution of gastroenteritis hospitalisations to norovirus in the winter. Therefore the reporting parameter linking infection to hospitalisation data varies between the winter and summer, and the transition between seasons is captured using a smooth cosine function.

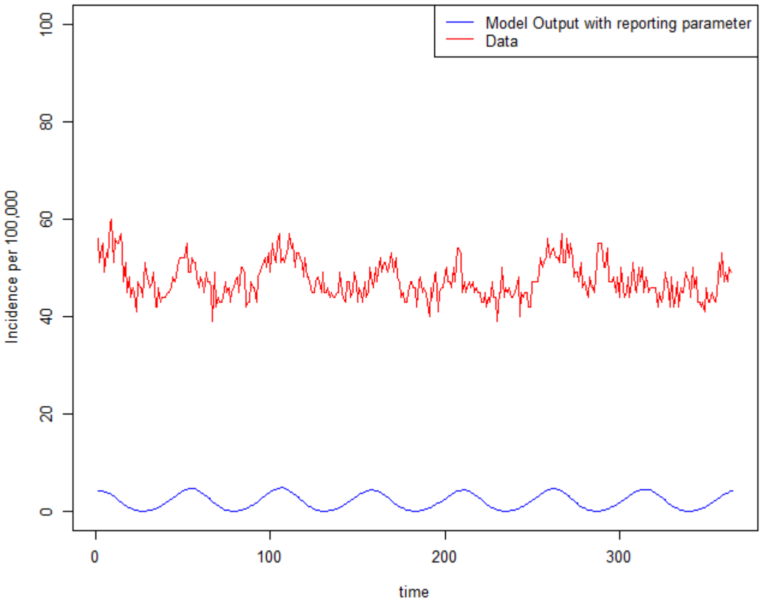

However, to fit the model output to the data on gastroenteritis hospitalisations, we need to incorporate the long term secular trend observed in the data and account for the other causes of gastroenteritis hospitalisations. First we capture the secular trend in the data with a B-spline basis function with 3 knots:

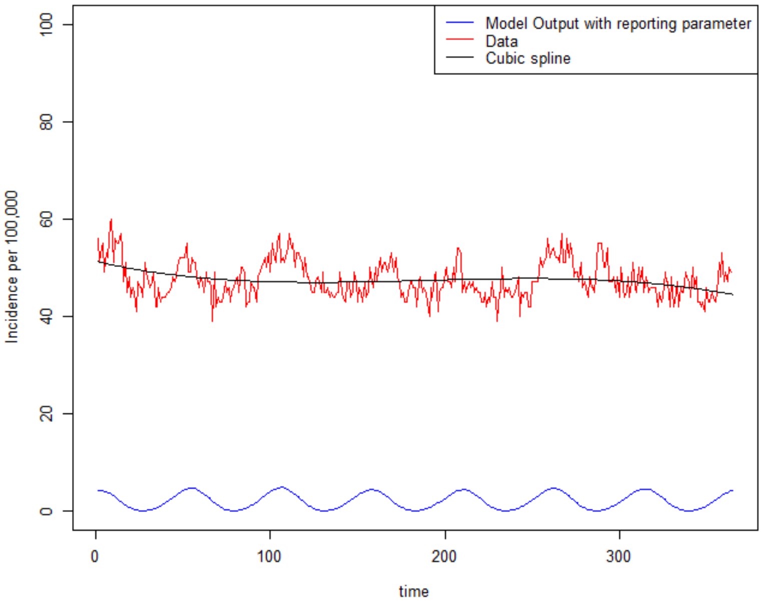

We then make an assumption that the sinusoidal seasonality in the gastroenteritis data is in part due to norovirus infections. In accounting for other causes of gastroenteritis hospitalisations, we simply add the changes in norovirus linked gastroenteritis hospitalisation incidence to the cubic spline (representing the secular trend of gastroenteritis hospitalisations, and other causes of gastroenteritis hospitalisations). The changes in the norovirus linked gastroenteritis hospitalisation incidence is calculated by taking the difference between the model output and the median of the model output to give an output as below.

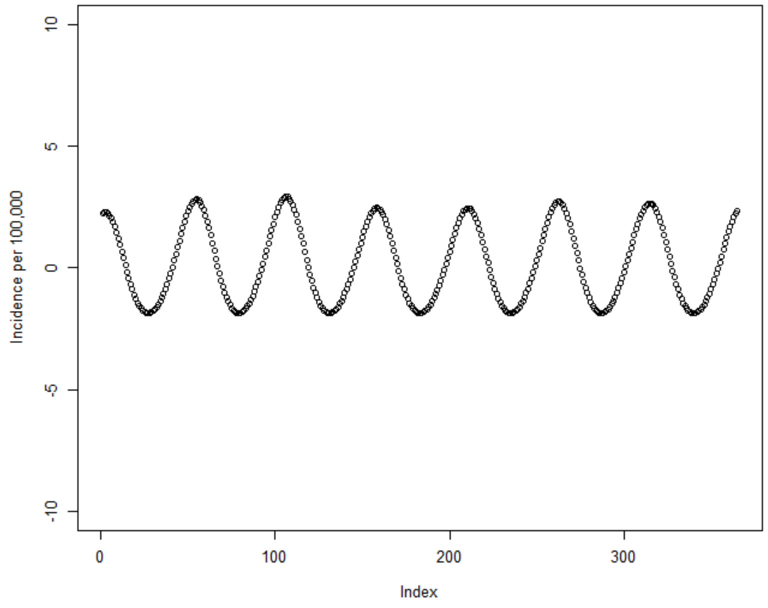

Then added to the cubic spline to fit to the data time series.

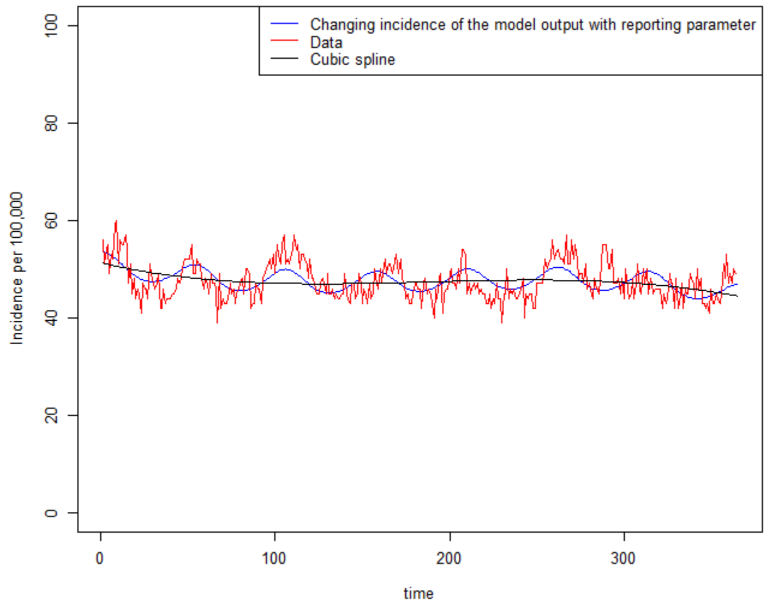

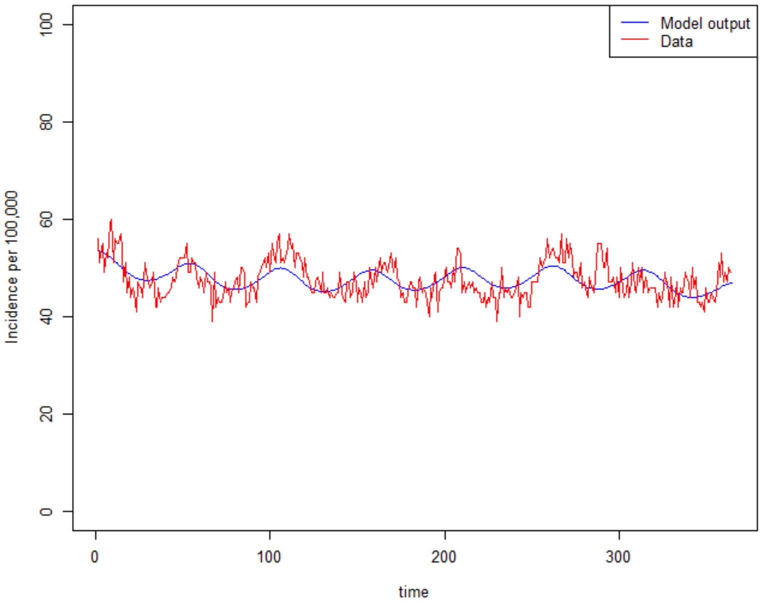

The parameter estimating the proportion of norovirus infections linked to gastroenteritis hospitalisations determines the amplitude of the model output fitting to the gastroenteritis hospitalisations data. Therefore assuming norovirus infections in part drives the sinusoidal seasonality observed in gastroenteritis hospitalisations, the larger proportion of norovirus linked AKI hospitalisation, then we would expect more extreme seasonality of the data.

#### S3.3 Observation model equations

##### S3.3.1 Fitting to surveillance data

*Number of notifications to surveillance stratified by age groups (0-4 and 15-64)*

$$A_i(t) = \theta_i \, Is_i(t)$$

*Number of notifications to surveillance for age group 65+*

$$A_{65+}(t) = Is_i \times \left[\left(\frac{1}{2}\left(1 + \cos\left(\frac{2\pi(w-1)}{52}\right)\right)\right) \times \theta_{winter,65+} + \left(\frac{1}{2}\left(1 - \cos\left(\frac{2\pi(w-1)}{52}\right)\right)\right) \times \theta_{summer,65+}\right]$$

Where:
- $A$ denotes the number of notifications to surveillance
- $i$ denotes age group (unless otherwise specified)
- $Is$ denotes the number of symptomatic infections simulated by the model
- $w$ denotes the week number
- $\theta_i$ denotes the reporting parameter scaling the number of infections by age group
- $\theta_{winter,65+}$ denotes the peak of the reporting parameter scaling the number of infections in the 65+ age group in the winter (week 52)
- $\theta_{summer,65+}$ denotes the trough of the reporting parameter scaling the number of infections in the 65+ age group in the summer (week 27)

##### S3.3.2 B-spline function

$$spl(t) = B_k(t)$$

Where:
- $k$ denotes the number of knots specified in the B-spline function
- $t$ is the variable at which the B-spline function is evaluated i.e. the function is evaluated at each time point

##### S3.3.3 Fitting to AKI hospitalisation data in 65+

$$Model\ incidence\ of\ AKI\ hospitalisation = B$$

$$spl_{AKI\ hosp,65+}(t) = (t_1, x_{1,AKI\ hosp}), (t_2, x_{2,AKI\ hosp)}), \ldots, (t_n, x_{n,AKI\ hosp}) \sim spl(t);$$

$$\theta_{AKI,seasonal,65+}(t) = Is_{65+} \times \left[\left(\frac{1}{2}\left(1 + \cos\left(\frac{2\pi(w-1)}{52}\right)\right)\right) \times \theta_{AKI,winter,65+} + \left(\frac{1}{2}\left(1 - \cos\left(\frac{2\pi(w-1)}{52}\right)\right)\right) \times \theta_{AKI,summer,65+}\right];$$

$$B(t) = \left(\left(\left(\frac{Is_{65+}}{pop_{65}} \times 100{,}000\right) \times \theta_{AKI,seasonal,65+}(t)\right) - median\left(\left(\frac{Is_{65+}}{pop_{65}} \times 100{,}000\right) \times \theta_{AKI,summer,65+}\right)\right) + spl_{AKI\ hosp,65+}(t)$$

Where:

- $B$ denotes the number of AKI hospitalisations (all cause)
- $t$ denotes time
- $spl_{AKI\ hosp,65+}$ denotes the spline function fitted to AKI hospitalisation data in 65+
- $\theta_{AKI,seasonal,65+}(t)$ denotes the seasonal reporting parameter, representing the proportion of norovirus infections linked to an AKI hospitalisation in a given time t
- $Is_{65+}$ denotes the number of symptomatic infections simulated by the model
- $w$ denotes the week number
- $\theta_{AKI,winter,65+}$ denotes the peak of the parameter scaling the number of infections in the 65+ age group in the winter (week 52) linked to an AKI hospitalisation in a given time t
- $\theta_{AKI,summer,65+}$ denotes the trough of the parameter scaling the number of infections in the 65+ age group in the summer (week 27) linked to an AKI hospitalisation in a given time t
- $pop_{65}$ denotes the population of the size of 65+ age group

#### S3.3.4 Fitting to gastroenteritis hospitalisation data in 65+

$$Model\ incidence\ of\ Gastroenteritis\ hospitalisation = C$$

$$spl_{Gastro\ hosp,65+}(t) = (t_1, x_{1,Gastro\ hosp}), (t_2, x_{2,Gastro\ hosp)}), \ldots, (t_3, x_{3,Gastro\ hosp}) \sim spl(t);$$

$$\theta_{Gastro,seasonal,65+}(t) = Is_{65+} \times \left[\left(\frac{1}{2}\left(1 + \cos\left(\frac{2\pi(w-1)}{52}\right)\right)\right) \times \theta_{Gastro,winter,65+} + \left(\frac{1}{2}\left(1 - \cos\left(\frac{2\pi(w-1)}{52}\right)\right)\right) \times \theta_{Gastro,summer,65+}\right];$$

$$C(t) = \left(\left(\left(\frac{Is_{65}}{pop_{65}} \times 100{,}000\right) \times \theta_{Gastro,seasonal,65+}(t)\right) - median\left(\left(\frac{Is_{65}}{pop_{65}} \times 100{,}000\right) \times \theta_{Gastro,summer,65+}\right)\right) + spl_{Gastro\ hosp,65+}(t)$$

Where:

- C denotes the number of gastroenteritis hospitalisations (all cause)
- $t$ denotes time
- $spl_{Gastro\ hosp,65+}$ denotes the spline function fitted to gastroenteritis hospitalisation data in 65+
- $\theta_{Gastro,seasonal,65+}(t)$ denotes the seasonal reporting parameter, representing the proportion of norovirus infections linked to a gastroenteritis hospitalisation in a given time t

- $Is_{65+}$ denotes the number of symptomatic infections simulated by the model
- $w$ denotes the week number
- $\theta_{Gastro,winter,65+}$ denotes the peak of the parameter scaling the number of infections in the 65+ age group in the winter (week 52) linked to a gastroenteritis hospitalisation in a given time t
- $\theta_{Gastro,summer,65+}$ denotes the trough of the parameter scaling the number of infections in the 65+ age group in the summer (week 27) linked to a gastroenteritis hospitalisation in a given time t
- $pop_{65}$ denotes the population size of 65+ age group

#### S3.3.5 Fitting to Gastroenteritis attendance to GP in 0-4

$$Model\ incidence\ of\ Gastroenteritis\ GP\ attendance\ in\ 0-4\ year\ olds\ = D$$

$$spl_{Gastro\ GP,0-4}(t) = (t_1, x_{1,Gastro\ GP}), (t_2, x_{2,Gastro\ GP}), \dots, (t_n, x_{n,Gastro\ GP}) \sim spl(t);$$

$$median\ spl_{Gastro\ GP,0-4}(t) = spl_{Gastro\ GP,0-4}(t) - median\left(spl_{Gastro\ GP,0-4}(t)\right);$$

$$D(t) = (\left(\frac{Is_{0-4}}{pop_{0-4}} \times 100{,}000\right) + median\ spl_{Gastro\ GP,0-4}(t)) \times \zeta$$

Where:

- D denotes the number gastroenteritis attendances in age group 0-4 (call cause)
- t denotes time
- $spl_{Gastro\ GP,0-4}$ denotes the spline function fitted to gastroenteritis GP attendances data in 0-4 age group
- $\zeta$ denotes the scaling parameter linking norovirus infections to the gastroenteritis GP attendances data in 0-4 age group
- $Is_{0-4}$ denotes the number of symptomatic infections simulated by the model
- $pop_{0-4}$ denotes the population size of 0-4 age group

#### S3.4 Likelihood calculation

To implement the Metropolis-Hastings algorithm we calculated an overall log likelihood for each value of θ. For each data source we calculated the likelihood assuming a quasi-Poisson distribution. For the age stratified incidence data a Poisson distribution was assumed. The log likelihood calculated for each source of data was then combined to give an overall log likelihood, which was used to estimate the posterior for each parameter (θ).

$$logL_{norovirus\ surveillance}\ (\theta | x, \phi_{noroivrus\ surveillance}) = \sum_{i=1}^{n} \left[x_i log(\mu_i(\theta)) - \mu_i(\theta) - log(x_i!)\right] / \phi_{noroivrus\ surveillance}$$

$$logL_{AKI\ hospitalisation}(\theta | x, \phi_{AKI\ hospitalisation}) = \sum_{i=1}^{n} \left[x_i log(\mu_i(\theta)) - \mu_i(\theta) - log(x_i!)\right] / \phi_{AKI\ hospitalisation}$$

$$logL_{Gastroenteritis\ hospitalisation}(\theta|x, \phi_{Gastroenteritis\ hospitalisation}) = \sum_{i=1}^{n}[x_i log(\mu_i(\theta)) - \mu_i(\theta) - log(x_i!)] \ / \ \phi_{Gastroenteritis\ hospitalisation}$$

$$logL_{Gastroenteritis\ GP\ attendance}(\theta|x, \phi_{Gastroenteritis\ GP\ attendance}) = \sum_{i=1}^{n}[x_i log(\mu_i(\theta)) - \mu_i(\theta) - log(x_i!)] \ / \ \phi_{Gastroenteritis\ GP\ attendance}$$

Where:

- L denotes the likelihood of each source of data
- θ denotes the parameters
- $x_i$ denotes the observed value of the *i-th* data point in each source of data
- $\mu_i$(θ) denotes the expected value for the *i-th* data point as a function of parameters θ
- $\phi$ denotes the dispersion parameter for each source of data

$$logL_{IID2\ age\ stratified\ incidence}(\theta|x) = \prod_{i=1}^{n} \frac{e^{-\theta}\theta^{x_i}}{x_i!}$$

Where:

- L denotes the likelihood
- θ denotes the parameters
- $x_i$ denotes the observed value of the *i*th data point

$$logL_{overall} = logL_{norovirus\ surveillance} + L_{AKI\ hospitalisation} + L_{Gastroenteritis\ hospitalisation} + L_{Gastroenteritis\ GP\ attendance} + L_{IID2\ age\ stratified\ incidence}$$

### S4. Descriptive analysis

#### S4.1 Age stratified norovirus surveillance time series

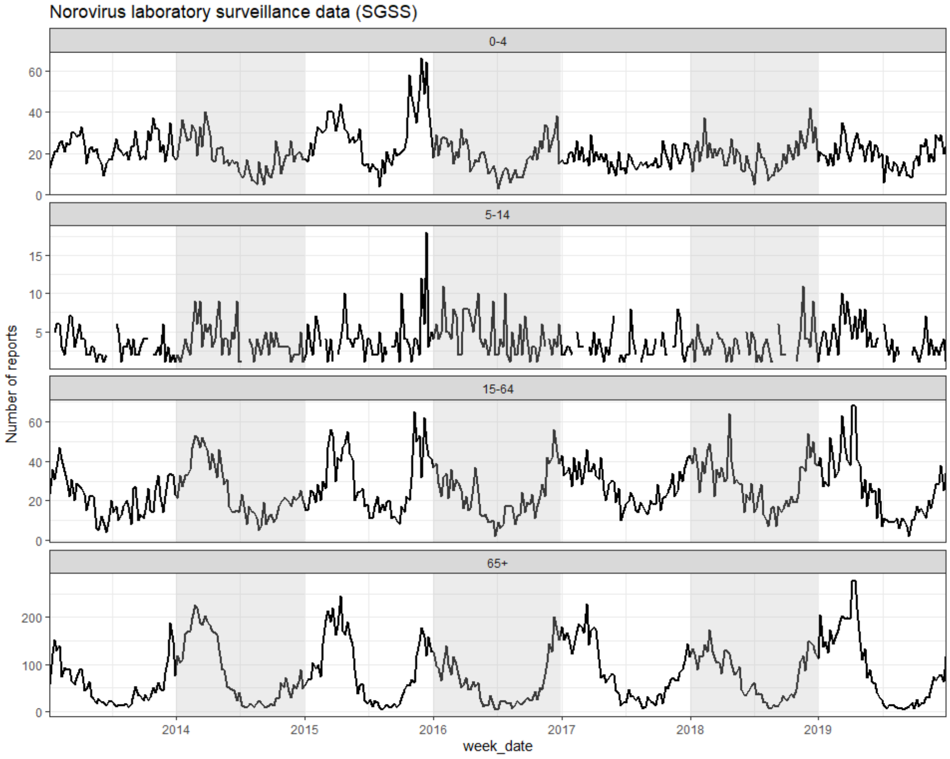

#### S4.2 Age stratified acute kidney injury hospitalisation time series

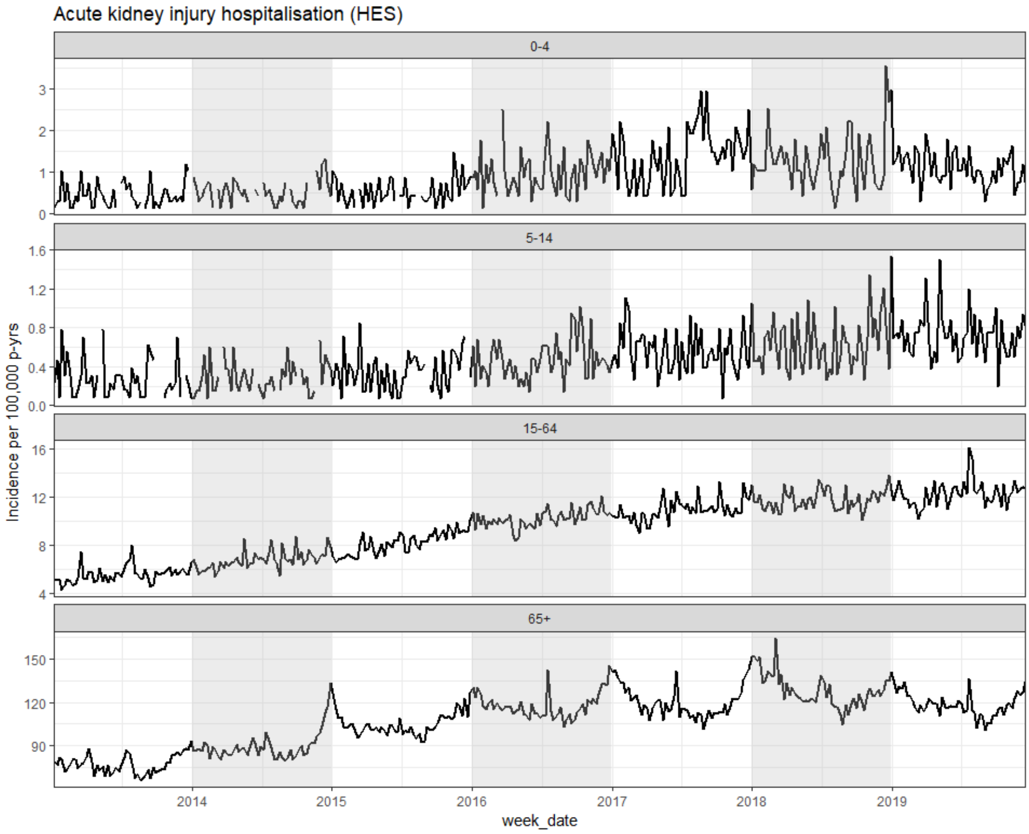

#### S4.3 Age stratified gastroenteritis hospitalisation time series

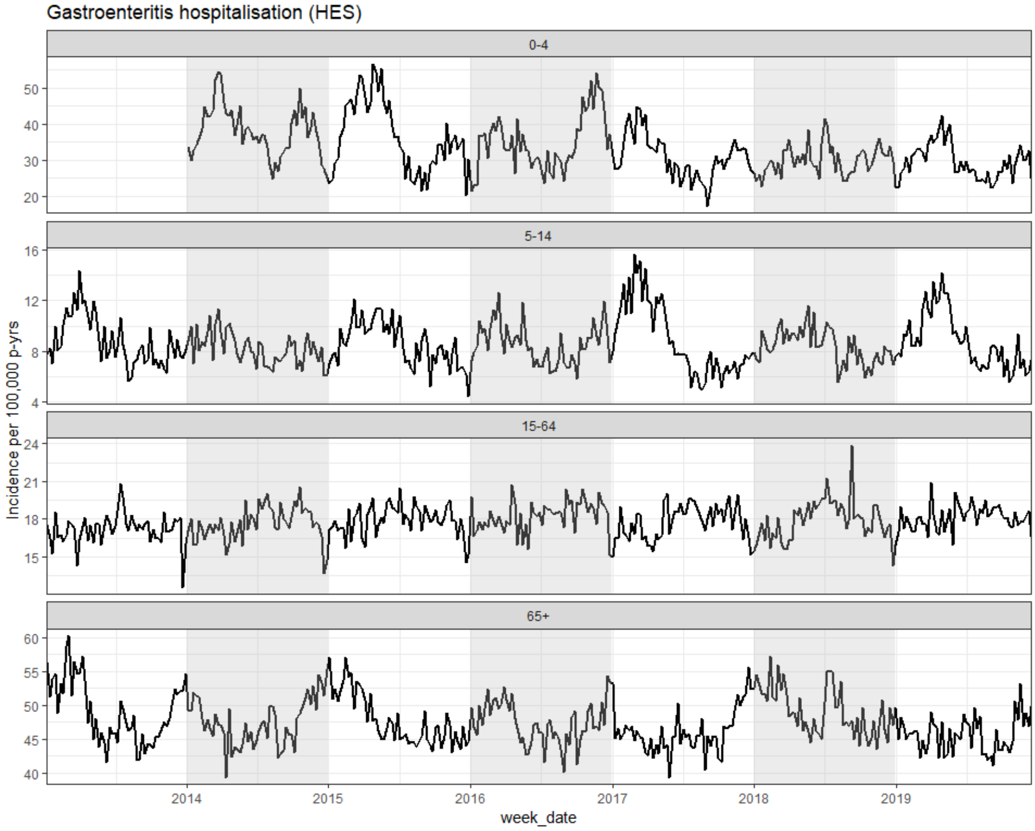

#### S4.4 Age stratified gastroenteritis GP attendance time series

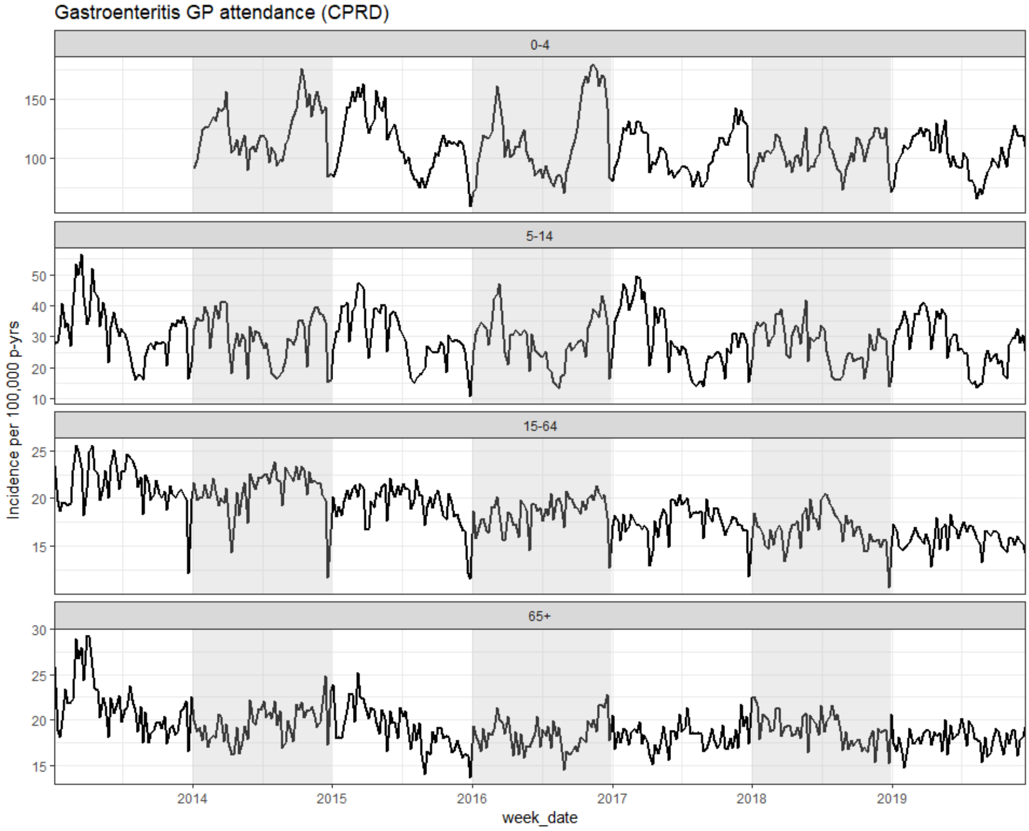

### S5. Assessment of model fit

#### S5.1 Trace plots of estimated parameters

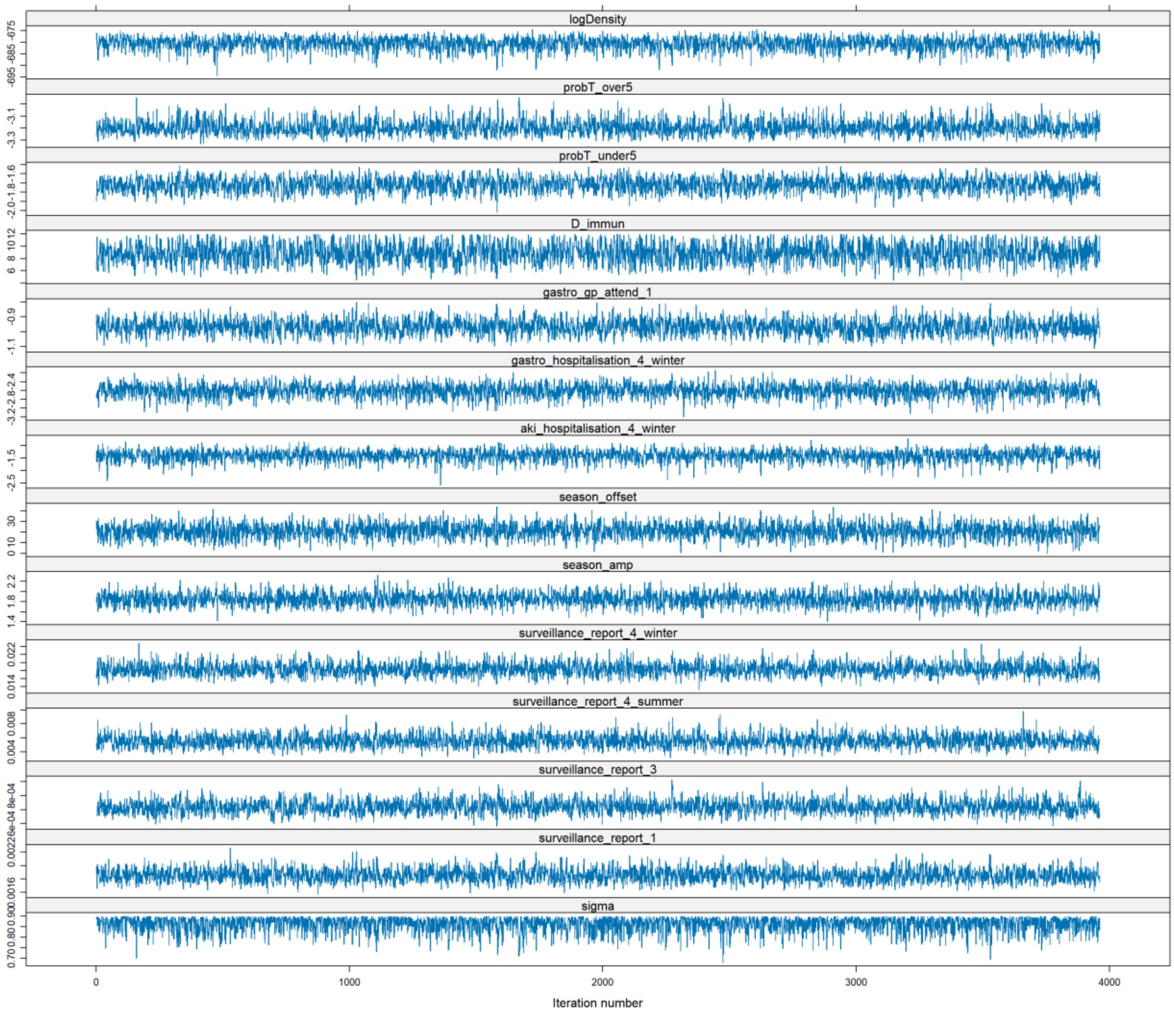

### S5.2 Posterior distribution of parameters

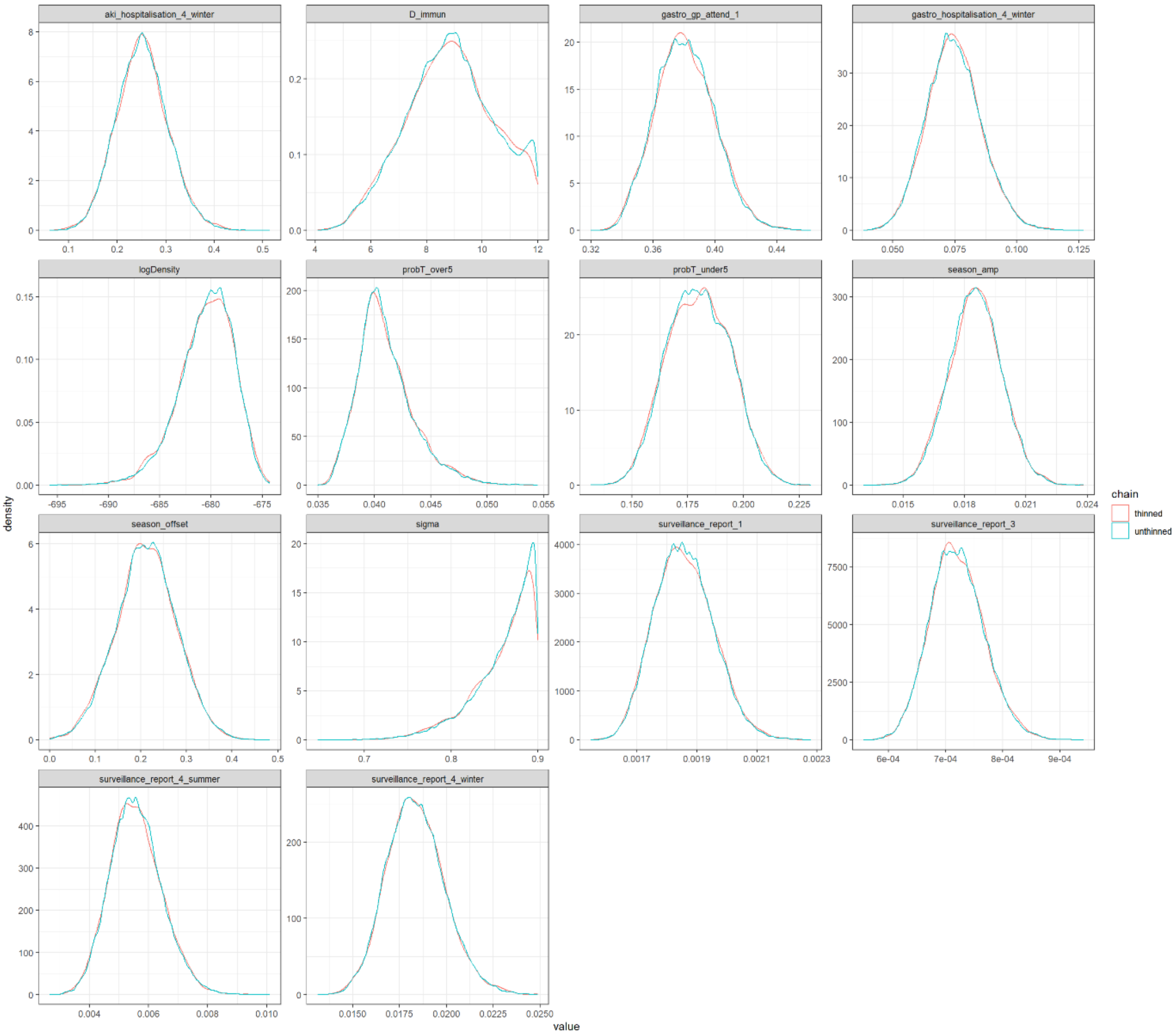

#### S5.3 Posterior and prior distributions

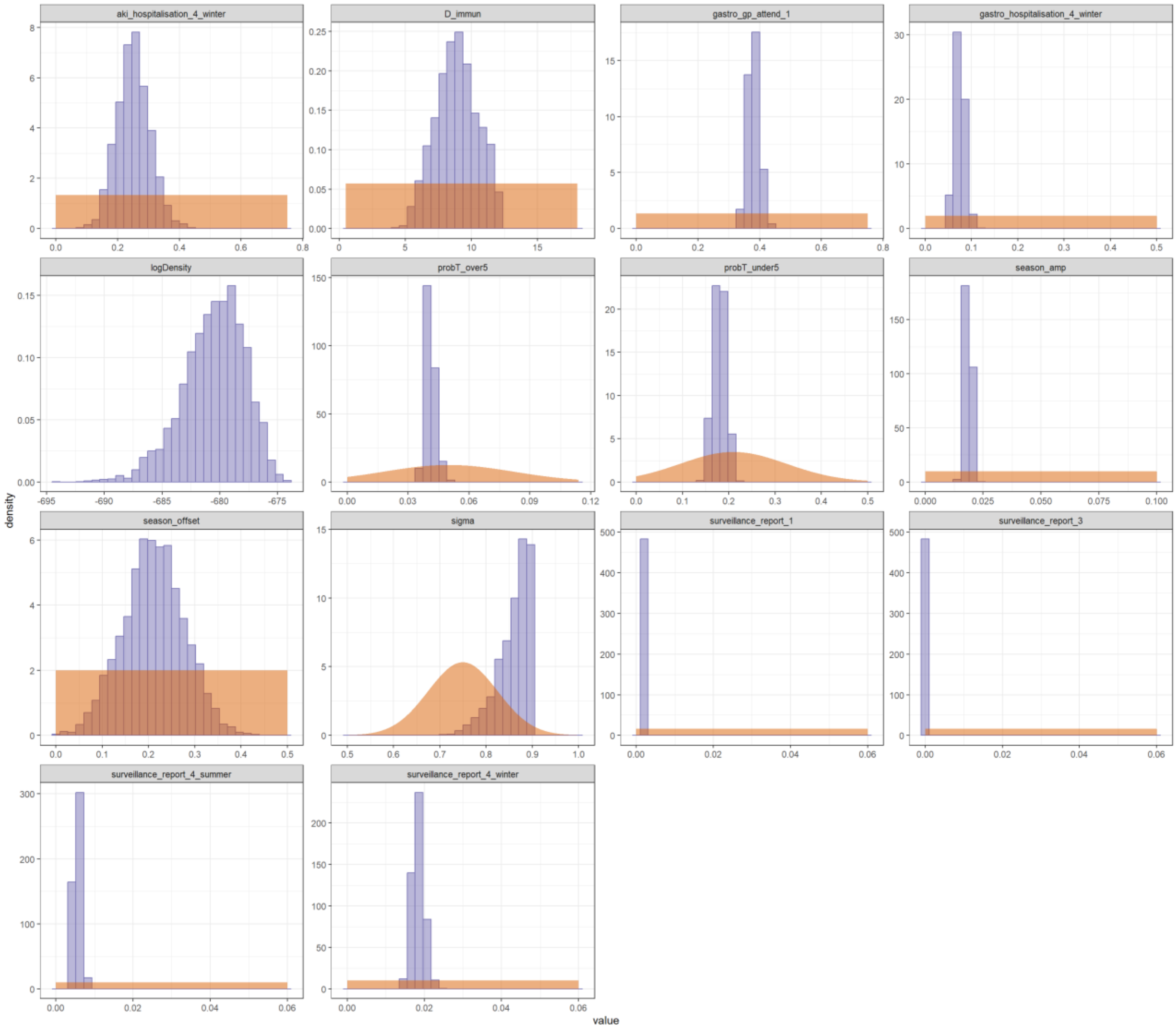

#### S5.4 Parameter correlations

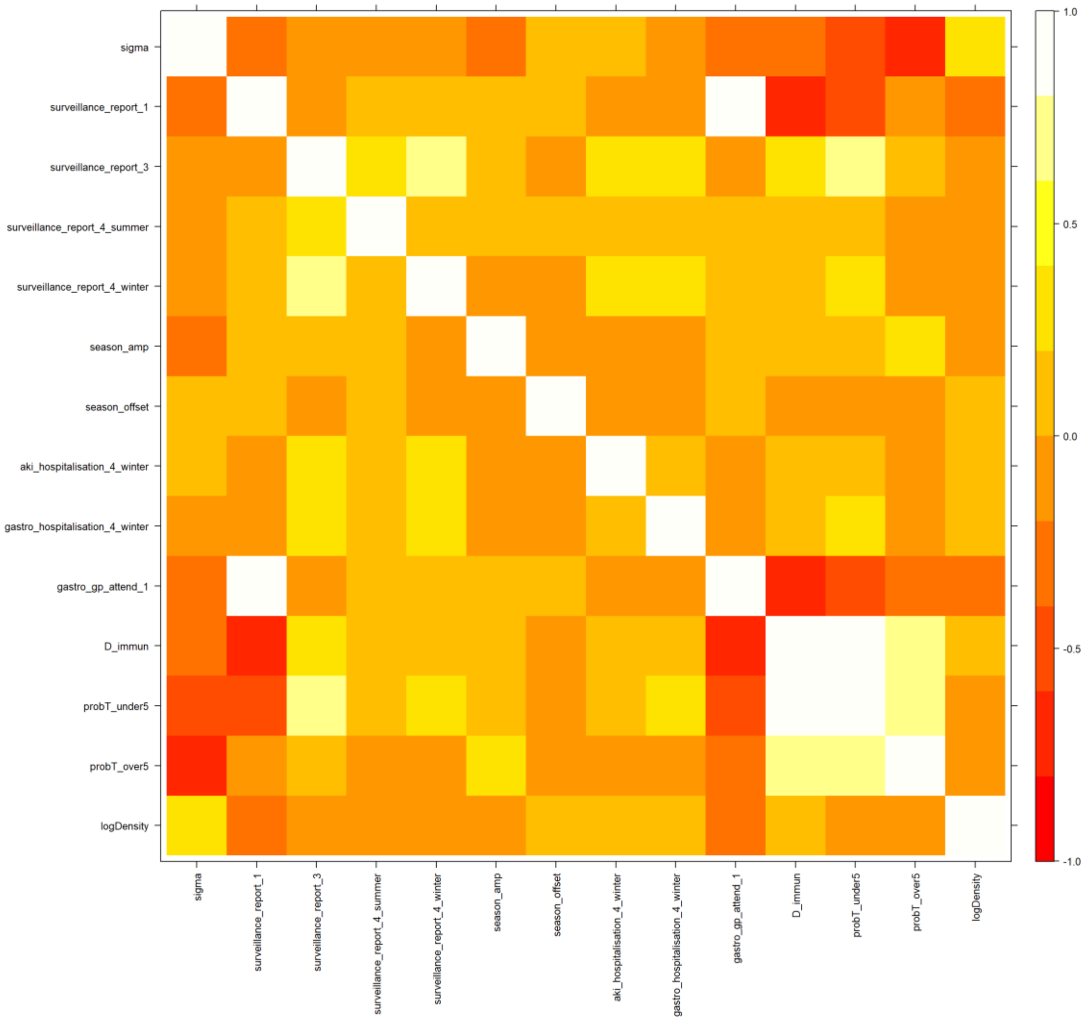

#### S5.5 Effective sample size

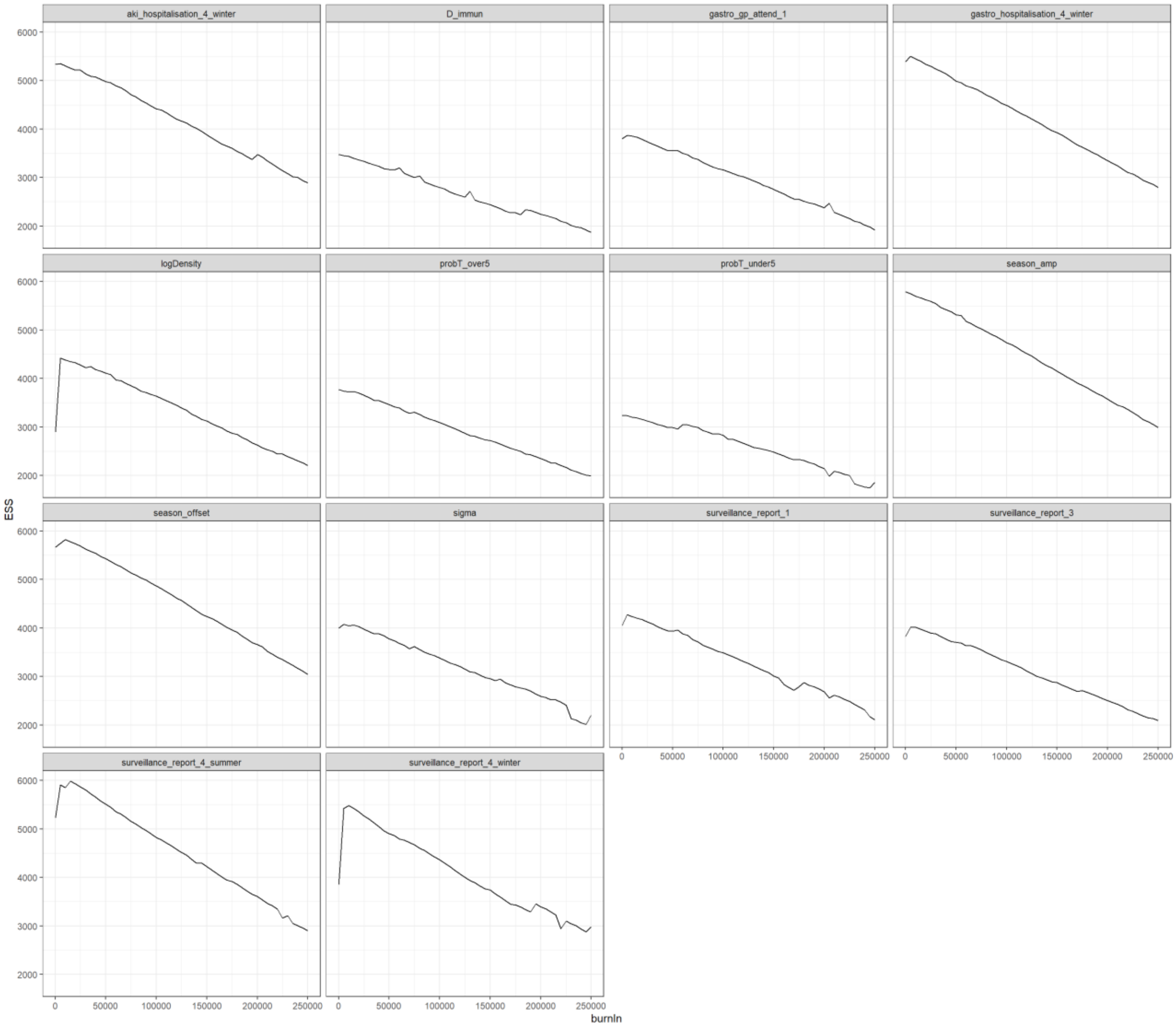

#### S5.6 Autocorrelation plots

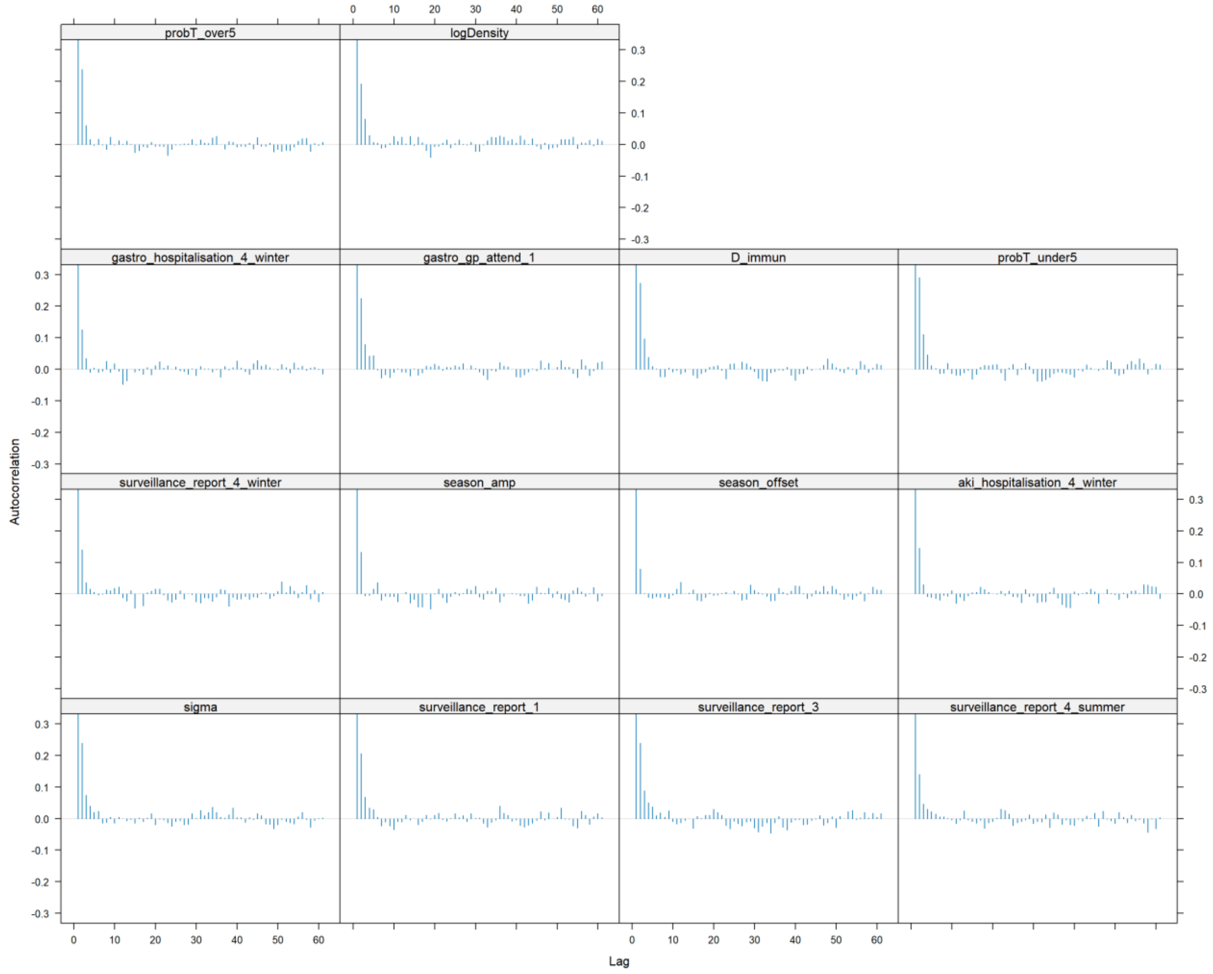

#### S5.7 Multi chain plots

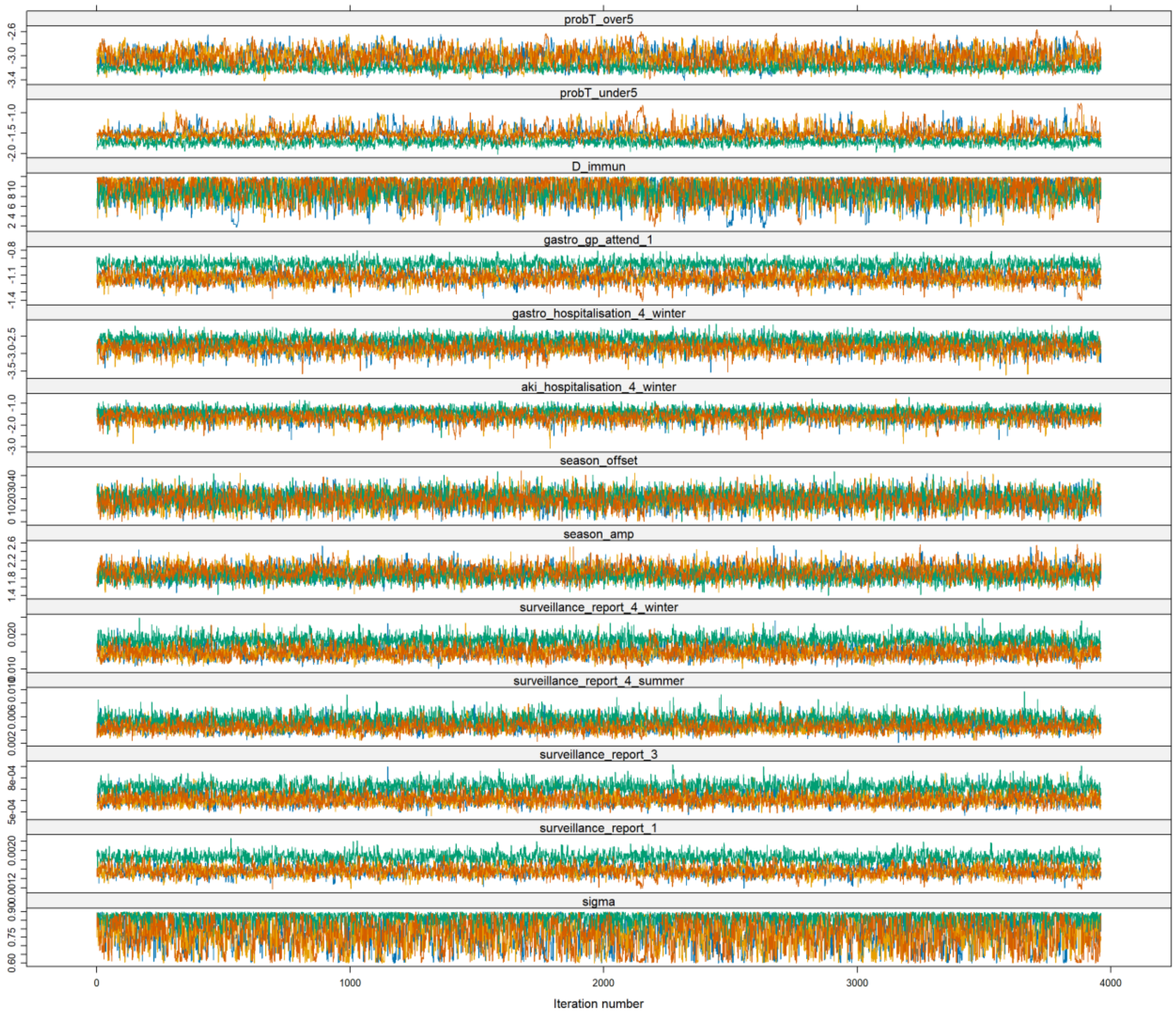

#### S5.8 Sensitivity analysis of AKI definition and spline knots

| Parameter | AKI definition - AKI code in primary and secondary position only | AKI definition - AKI code any positon and any time during admission | AKI definition 1 knot | AKI definition 2 knots |
|---|---|---|---|---|
| Proportion symptomatic | 0.78 (0.61-0.89) | 0.8 (0.62-0.89) | 0.87 (0.76-0.9) | 0.78 (0.62-0.89) |
| Duration of immunity | 9.7 (4.2-12) | 10 (5.1-12) | 8.9 (5.9-12) | 10 (5.2-12) |
| Probability of infection between under 5s | 0.22 (0.18-0.31) | 0.22 (0.18-0.3) | 0.18 (0.16-0.21) | 0.22 (0.18-0.3) |
| Probability of infection to over 5s | 0.048 (0.038-0.064) | 0.048 (0.039-0.062) | 0.041 (0.037-0.048) | 0.049 (0.039-0.061) |
| Seasonal amplitude term | 0.019 (0.016-0.023) | 0.02 (0.017-0.023) | 0.019 (0.016-0.021) | 0.02 (0.017-0.022) |
| Seasonal offset term | 0.19 (0.06-0.31) | 0.2 (0.067-0.34) | 0.22 (0.082-0.34) | 0.19 (0.057-0.32) |
| Norovirus associated AKI hospitalisation in 65+ in the winter | 0.0023 (0.0011-0.014) | 0.23 (0.14-0.32) | 0.26 (0.17-0.37) | 0.2 (0.13-0.3) |
| Norovirus associated hospitalisation in 65+ in the winter | 0.065 (0.048-0.088) | 0.063 (0.048-0.083) | 0.08 (0.059-0.1) | 0.063 (0.047-0.087) |
| Underreporting to surveillance 0-4 | 0.0016 (0.0014-0.0018) | 0.0016 (0.0014-0.0018) | 0.0019 (0.0017-0.002) | 0.0015 (0.0014-0.0018) |
| Underreporting to surveillance 15-64 | 0.00062 (0.00053-0.00072) | 0.00061 (0.00053-0.00072) | 0.00072 (0.00064-0.00083) | 6e-04 (0.00053-0.00071) |
| Underreporting to surveillance 65+ in the winter | 0.015 (0.012-0.019) | 0.015 (0.012-0.019) | 0.018 (0.016-0.022) | 0.015 (0.012-0.018) |
| Underreporting to surveillance 65+ in the summer | 0.0045 (0.0032-0.0063) | 0.0045 (0.0031-0.0064) | 0.0055 (0.004-0.0074) | 0.0044 (0.0031-0.0062) |
| GP attendance for all cause gastroenteritis in under 5s | 0.32 (0.29-0.37) | 0.32 (0.29-0.37) | 0.38 (0.35-0.42) | 0.32 (0.29-0.37) |

### S6. Cost analysis

#### S6.1 Elective inpatient and non-elective inpatient costs (21,22)

| currency | currency_description | activity | unit_cost | total_cost | year | type |
|---|---|---|---|---|---|---|
| LA07H | Acute Kidney Injury with Interventions, with CC Score 11+ | 17 | 7,907.799 | 134,432.58 | 2013-2014 | Elective Inpatients |
| LA07J | Acute Kidney Injury with Interventions, with CC Score 6-10 | 59 | 7,241.908 | 427,272.57 | 2013-2014 | Elective Inpatients |
| LA07K | Acute Kidney Injury with Interventions, with CC Score 0-5 | 104 | 4,603.212 | 478,734.05 | 2013-2014 | Elective Inpatients |
| LA07L | Acute Kidney Injury without Interventions, with CC Score 12+ | 34 | 4,168.357 | 141,724.14 | 2013-2014 | Elective Inpatients |
| LA07M | Acute Kidney Injury without Interventions, with CC Score 8-11 | 116 | 3,275.483 | 379,956.00 | 2013-2014 | Elective Inpatients |
| LA07N | Acute Kidney Injury without Interventions, with CC Score 4-7 | 310 | 2,585.044 | 801,363.60 | 2013-2014 | Elective Inpatients |
| LA07P | Acute Kidney Injury without Interventions, with CC Score 0-3 | 536 | 1,696.352 | 909,244.85 | 2013-2014 | Elective Inpatients |
| LA07H | Acute Kidney Injury with Interventions, with CC Score 11+ | 18 | 7,902.517 | 142,245.30 | 2014-2015 | Elective Inpatients |
| LA07J | Acute Kidney Injury with Interventions, with CC Score 6-10 | 78 | 6,749.594 | 526,468.31 | 2014-2015 | Elective Inpatients |
| LA07K | Acute Kidney Injury with Interventions, with CC Score 0-5 | 175 | 3,869.496 | 677,161.87 | 2014-2015 | Elective Inpatients |
| LA07L | Acute Kidney Injury without Interventions, with CC Score 12+ | 47 | 4,269.758 | 200,678.62 | 2014-2015 | Elective Inpatients |
| LA07M | Acute Kidney Injury without Interventions, with CC Score 8-11 | 127 | 2,532.303 | 321,602.43 | 2014-2015 | Elective Inpatients |
| LA07N | Acute Kidney Injury without Interventions, with CC Score 4-7 | 405 | 2,233.162 | 904,430.44 | 2014-2015 | Elective Inpatients |
| LA07P | Acute Kidney Injury without Interventions, with CC Score 0-3 | 616 | 1,402.640 | 864,026.21 | 2014-2015 | Elective Inpatients |
| LA07H | Acute Kidney Injury with Interventions, with CC Score 11+ | 31 | 9,579.441 | 296,962.68 | 2015-2016 | Elective Inpatients |
| LA07J | Acute Kidney Injury with Interventions, with CC Score 6-10 | 92 | 7,569.423 | 696,386.94 | 2015-2016 | Elective Inpatients |
| LA07K | Acute Kidney Injury with Interventions, with CC Score 0-5 | 191 | 3,601.290 | 687,846.30 | 2015-2016 | Elective Inpatients |
| LA07L | Acute Kidney Injury without Interventions, with CC Score 12+ | 27 | 3,354.564 | 90,573.24 | 2015-2016 | Elective Inpatients |
| LA07M | Acute Kidney Injury without Interventions, with CC Score 8-11 | 159 | 3,373.126 | 536,327.00 | 2015-2016 | Elective Inpatients |
| LA07N | Acute Kidney Injury without Interventions, with CC Score 4-7 | 510 | 2,087.279 | 1,064,512.47 | 2015-2016 | Elective Inpatients |
| LA07P | Acute Kidney Injury without Interventions, with CC Score 0-3 | 714 | 1,493.748 | 1,066,536.30 | 2015-2016 | Elective Inpatients |
| LA07H | Acute Kidney Injury with Interventions, with CC Score 11+ | 17 | 8,612.242 | 146,408.12 | 2016-2017 | Elective Inpatients |
| LA07J | Acute Kidney Injury with Interventions, with CC Score 6-10 | 89 | 5,889.121 | 524,131.80 | 2016-2017 | Elective Inpatients |
| LA07K | Acute Kidney Injury with Interventions, with CC Score 0-5 | 212 | 3,956.070 | 838,686.89 | 2016-2017 | Elective Inpatients |
| LA07L | Acute Kidney Injury without Interventions, with CC Score 12+ | 39 | 3,969.871 | 154,824.95 | 2016-2017 | Elective Inpatients |
| LA07M | Acute Kidney Injury without Interventions, with CC Score 8-11 | 161 | 2,874.770 | 462,837.89 | 2016-2017 | Elective Inpatients |
| LA07N | Acute Kidney Injury without Interventions, with CC Score 4-7 | 487 | 1,766.250 | 860,163.74 | 2016-2017 | Elective Inpatients |
| LA07P | Acute Kidney Injury without Interventions, with CC Score 0-3 | 792 | 1,359.246 | 1,076,522.95 | 2016-2017 | Elective Inpatients |
| LA07H | Acute Kidney Injury with Interventions, with CC Score 11+ | 30 | 8,451.193 | 253,535.79 | 2017-2018 | Elective Inpatients |
| LA07J | Acute Kidney Injury with Interventions, with CC Score 6-10 | 107 | 5,647.583 | 604,291.42 | 2017-2018 | Elective Inpatients |
| LA07K | Acute Kidney Injury with Interventions, with CC Score 0-5 | 214 | 4,144.560 | 886,935.78 | 2017-2018 | Elective Inpatients |
| LA07L | Acute Kidney Injury without Interventions, with CC Score 12+ | 34 | 4,432.709 | 150,712.09 | 2017-2018 | Elective Inpatients |
| LA07M | Acute Kidney Injury without Interventions, with CC Score 8-11 | 228 | 2,566.488 | 585,159.15 | 2017-2018 | Elective Inpatients |
| LA07N | Acute Kidney Injury without Interventions, with CC Score 4-7 | 571 | 1,773.467 | 1,012,649.40 | 2017-2018 | Elective Inpatients |
| LA07P | Acute Kidney Injury without Interventions, with CC Score 0-3 | 658 | 1,338.331 | 880,621.63 | 2017-2018 | Elective Inpatients |
| LA07H | Acute Kidney Injury with Interventions, with CC Score 11+ | 41 | 9,189.626 | 376,774.68 | 2018-2019 | Elective Inpatients |
| LA07J | Acute Kidney Injury with Interventions, with CC Score 6-10 | 95 | 5,820.393 | 552,937.31 | 2018-2019 | Elective Inpatients |
| LA07K | Acute Kidney Injury with Interventions, with CC Score 0-5 | 216 | 3,879.436 | 837,958.10 | 2018-2019 | Elective Inpatients |
| LA07L | Acute Kidney Injury without Interventions, with CC Score 12+ | 50 | 5,637.573 | 281,878.67 | 2018-2019 | Elective Inpatients |
| LA07M | Acute Kidney Injury without Interventions, with CC Score 8-11 | 227 | 3,284.832 | 745,656.87 | 2018-2019 | Elective Inpatients |

| currency | currency_description | activity | unit_cost | total_cost | year | type |
|---|---|---|---|---|---|---|
| LA07N | Acute Kidney Injury without Interventions, with CC Score 4-7 | 527 | 1,879.562 | 990,529.08 | 2018-2019 | Elective Inpatients |
| LA07P | Acute Kidney Injury without Interventions, with CC Score 0-3 | 673 | 1,476.884 | 993,942.87 | 2018-2019 | Elective Inpatients |
| LA07H | Acute Kidney Injury with Interventions, with CC Score 11+ | 895 | 6,433.694 | 5,758,156.50 | 2013-2014 | Non-Elective Inpatients - Long Stay |
| LA07J | Acute Kidney Injury with Interventions, with CC Score 6-10 | 2,597 | 4,812.611 | 12,498,349.94 | 2013-2014 | Non-Elective Inpatients - Long Stay |
| LA07K | Acute Kidney Injury with Interventions, with CC Score 0-5 | 2,194 | 3,512.228 | 7,705,828.44 | 2013-2014 | Non-Elective Inpatients - Long Stay |
| LA07L | Acute Kidney Injury without Interventions, with CC Score 12+ | 2,024 | 3,663.807 | 7,415,544.72 | 2013-2014 | Non-Elective Inpatients - Long Stay |
| LA07M | Acute Kidney Injury without Interventions, with CC Score 8-11 | 8,304 | 2,891.999 | 24,015,163.76 | 2013-2014 | Non-Elective Inpatients - Long Stay |
| LA07N | Acute Kidney Injury without Interventions, with CC Score 4-7 | 17,213 | 2,224.495 | 38,290,238.27 | 2013-2014 | Non-Elective Inpatients - Long Stay |
| LA07P | Acute Kidney Injury without Interventions, with CC Score 0-3 | 10,939 | 1,746.525 | 19,105,234.75 | 2013-2014 | Non-Elective Inpatients - Long Stay |
| LA07H | Acute Kidney Injury with Interventions, with CC Score 11+ | 1,062 | 6,587.701 | 6,996,138.57 | 2014-2015 | Non-Elective Inpatients - Long Stay |
| LA07J | Acute Kidney Injury with Interventions, with CC Score 6-10 | 3,140 | 5,016.299 | 15,751,179.57 | 2014-2015 | Non-Elective Inpatients - Long Stay |
| LA07K | Acute Kidney Injury with Interventions, with CC Score 0-5 | 3,059 | 3,784.718 | 11,577,452.98 | 2014-2015 | Non-Elective Inpatients - Long Stay |
| LA07L | Acute Kidney Injury without Interventions, with CC Score 12+ | 2,343 | 3,786.669 | 8,872,166.49 | 2014-2015 | Non-Elective Inpatients - Long Stay |
| LA07M | Acute Kidney Injury without Interventions, with CC Score 8-11 | 9,321 | 2,953.558 | 27,530,112.62 | 2014-2015 | Non-Elective Inpatients - Long Stay |
| LA07N | Acute Kidney Injury without Interventions, with CC Score 4-7 | 19,564 | 2,227.329 | 43,575,473.89 | 2014-2015 | Non-Elective Inpatients - Long Stay |
| LA07P | Acute Kidney Injury without Interventions, with CC Score 0-3 | 11,622 | 1,785.630 | 20,752,588.57 | 2014-2015 | Non-Elective Inpatients - Long Stay |
| LA07H | Acute Kidney Injury with Interventions, with CC Score 11+ | 1,221 | 6,720.552 | 8,205,794.33 | 2015-2016 | Non-Elective Inpatients - Long Stay |
| LA07J | Acute Kidney Injury with Interventions, with CC Score 6-10 | 3,580 | 4,995.541 | 17,884,034.99 | 2015-2016 | Non-Elective Inpatients - Long Stay |
| LA07K | Acute Kidney Injury with Interventions, with CC Score 0-5 | 3,220 | 3,779.535 | 12,170,102.11 | 2015-2016 | Non-Elective Inpatients - Long Stay |
| LA07L | Acute Kidney Injury without Interventions, with CC Score 12+ | 2,725 | 3,871.695 | 10,550,369.31 | 2015-2016 | Non-Elective Inpatients - Long Stay |
| LA07M | Acute Kidney Injury without Interventions, with CC Score 8-11 | 10,601 | 2,893.770 | 30,676,855.83 | 2015-2016 | Non-Elective Inpatients - Long Stay |
| LA07N | Acute Kidney Injury without Interventions, with CC Score 4-7 | 20,202 | 2,163.854 | 43,714,177.99 | 2015-2016 | Non-Elective Inpatients - Long Stay |
| LA07P | Acute Kidney Injury without Interventions, with CC Score 0-3 | 11,837 | 1,812.846 | 21,458,652.46 | 2015-2016 | Non-Elective Inpatients - Long Stay |
| LA07H | Acute Kidney Injury with Interventions, with CC Score 11+ | 1,365 | 6,714.072 | 9,164,708.20 | 2016-2017 | Non-Elective Inpatients - Long Stay |
| LA07J | Acute Kidney Injury with Interventions, with CC Score 6-10 | 3,817 | 4,670.266 | 17,826,406.75 | 2016-2017 | Non-Elective Inpatients - Long Stay |
| LA07K | Acute Kidney Injury with Interventions, with CC Score 0-5 | 3,304 | 3,605.986 | 11,914,178.21 | 2016-2017 | Non-Elective Inpatients - Long Stay |
| LA07L | Acute Kidney Injury without Interventions, with CC Score 12+ | 3,184 | 3,608.385 | 11,489,096.70 | 2016-2017 | Non-Elective Inpatients - Long Stay |
| LA07M | Acute Kidney Injury without Interventions, with CC Score 8-11 | 11,257 | 2,751.746 | 30,976,403.09 | 2016-2017 | Non-Elective Inpatients - Long Stay |
| LA07N | Acute Kidney Injury without Interventions, with CC Score 4-7 | 20,236 | 2,107.993 | 42,657,350.23 | 2016-2017 | Non-Elective Inpatients - Long Stay |
| LA07P | Acute Kidney Injury without Interventions, with CC Score 0-3 | 11,525 | 1,662.109 | 19,155,807.55 | 2016-2017 | Non-Elective Inpatients - Long Stay |
| LA07H | Acute Kidney Injury with Interventions, with CC Score 11+ | 1,679 | 6,294.697 | 10,568,796.64 | 2017-2018 | Non-Elective Inpatients - Long Stay |
| LA07J | Acute Kidney Injury with Interventions, with CC Score 6-10 | 3,417 | 4,609.432 | 15,750,430.18 | 2017-2018 | Non-Elective Inpatients - Long Stay |
| LA07K | Acute Kidney Injury with Interventions, with CC Score 0-5 | 2,960 | 3,581.195 | 10,600,335.91 | 2017-2018 | Non-Elective Inpatients - Long Stay |
| LA07L | Acute Kidney Injury without Interventions, with CC Score 12+ | 4,424 | 3,564.853 | 15,770,908.05 | 2017-2018 | Non-Elective Inpatients - Long Stay |
| LA07M | Acute Kidney Injury without Interventions, with CC Score 8-11 | 11,354 | 2,703.707 | 30,697,884.53 | 2017-2018 | Non-Elective Inpatients - Long Stay |
| LA07N | Acute Kidney Injury without Interventions, with CC Score 4-7 | 18,525 | 2,074.279 | 38,426,016.52 | 2017-2018 | Non-Elective Inpatients - Long Stay |
| LA07P | Acute Kidney Injury without Interventions, with CC Score 0-3 | 9,889 | 1,808.682 | 17,886,054.36 | 2017-2018 | Non-Elective Inpatients - Long Stay |
| LA07H | Acute Kidney Injury with Interventions, with CC Score 11+ | 2,015 | 5,655.508 | 11,395,847.69 | 2018-2019 | Non-Elective Inpatients - Long Stay |
| LA07J | Acute Kidney Injury with Interventions, with CC Score 6-10 | 3,533 | 4,695.233 | 16,588,259.37 | 2018-2019 | Non-Elective Inpatients - Long Stay |
| LA07K | Acute Kidney Injury with Interventions, with CC Score 0-5 | 2,724 | 3,692.976 | 10,059,667.71 | 2018-2019 | Non-Elective Inpatients - Long Stay |
| LA07L | Acute Kidney Injury without Interventions, with CC Score 12+ | 5,666 | 3,512.503 | 19,901,842.32 | 2018-2019 | Non-Elective Inpatients - Long Stay |
| LA07M | Acute Kidney Injury without Interventions, with CC Score 8-11 | 12,380 | 2,854.320 | 35,336,485.08 | 2018-2019 | Non-Elective Inpatients - Long Stay |
| LA07N | Acute Kidney Injury without Interventions, with CC Score 4-7 | 18,948 | 2,313.415 | 43,834,581.94 | 2018-2019 | Non-Elective Inpatients - Long Stay |
| LA07P | Acute Kidney Injury without Interventions, with CC Score 0-3 | 10,071 | 1,955.560 | 19,694,441.42 | 2018-2019 | Non-Elective Inpatients - Long Stay |

### S7. Code list

#### S7.1 ICD-10 codes

| ICD10 code | Code description |
|---|---|
| N17.0 | Acute renal failure with tubular necrosis |
| N17.1 | Acute renal failure with acute cortical necrosis |
| N17.2 | Acute renal failure with medullary necrosis |
| N17.8 | Other acute renal failure |
| N17.9 | Acute renal failure, unspecified |
| N19 | Unspecified kidney failure |
| A00.0 | Cholera due to Vibrio cholerae 01, biovar cholerae |
| A00.1 | Cholera due to Vibrio cholerae 01, biovar eltor |
| A00.9 | Cholera, unspecified |
| A01.0 | Typhoid fever |
| A01.1 | Paratyphoid fever A |
| A01.2 | Paratyphoid fever B |
| A01.3 | Paratyphoid fever C |
| A01.4 | Paratyphoid fever, unspecified |
| A02.0 | Salmonella enteritis |
| A02.1 | Salmonella sepsis |
| A02.2 | Localized salmonella infections |
| A02.8 | Other specified salmonella infections |
| A02.9 | Salmonella infection, unspecified |
| A03.0 | Shigellosis due to Shigella dysenteriae |
| A03.1 | Shigellosis due to Shigella flexneri |
| A03.2 | Shigellosis due to Shigella boydii |
| A03.3 | Shigellosis due to Shigella sonnei |
| A03.8 | Other shigellosis |
| A03.9 | Shigellosis, unspecified |
| A04.0 | Enteropathogenic Escherichia coli infection |
| A04.1 | Enterotoxigenic Escherichia coli infection |
| A04.2 | Enteroinvasive Escherichia coli infection |
| A04.3 | Enterohaemorrhagic Escherichia coli infection |
| A04.4 | Other intestinal Escherichia coli infections |
| A04.5 | Campylobacter enteritis |
| A04.6 | Enteritis due to Yersinia enterocolitica |
| A04.7 | Enterocolitis due to Clostridium difficile |
| A04.8 | Other specified bacterial intestinal infections |
| A04.9 | Bacterial intestinal infection, unspecified |
| A05.0 | Other bacterial foodborne intoxications, not elsewhere classified |
| A05.1 | Botulism |
| A05.2 | Foodborne Clostridium perfringens [Clostridium welchii] intoxication |
| A05.3 | Foodborne Vibrio parahaemolyticus intoxication |
| A05.4 | Foodborne Bacillus cereus intoxication |
| A05.8 | Other specified bacterial foodborne intoxications |
| A05.9 | Bacterial foodborne intoxication, unspecified |
| A06.0 | Acute amoebic dysentery |
| A06.1 | Chronic intestinal amoebiasis |
| A06.2 | Amoebic nondysenteric colitis |
| A06.3 | Amoeboma of intestine |
| A06.4 | Amoebic liver abscess (K77.0*) |
| A06.5 | Amoebic lung abscess (J99.8*) |
| A06.6 | Amoebic brain abscess (G07*) |
| A06.7 | Cutaneous amoebiasis |
| A06.8 | Amoebic infection of other sites |
| A06.9 | Amoebiasis, unspecified |
| A07.0 | Balantidiasis |
| A07.1 | Giardiasis [lambliasis] |
| A07.2 | Cryptosporidiosis |
| A07.3 | Isosporiasis |
| A07.8 | Other specified protozoal intestinal diseases |
| A07.9 | Protozoal intestinal disease, unspecified |
| A08.0 | Rotaviral enteritis |
| A08.1 | Acute gastroenteropathy due to Norwalk agent |
| A08.2 | Adenoviral enteritis |
| A08.3 | Other viral enteritis |
| A08.4 | Viral intestinal infection, unspecified |
| A08.5 | Other specified intestinal infections |
| A09 | Other gastroenteritis and colitis of infectious and unspecified origin |
| A09.0 | Other and unspecified gastroenteritis and colitis of infectious origin |
| A09.9 | Gastroenteritis and colitis of unspecified origin |

| ICD10 code | Code description |
|---|---|
| K52.8 | Other specified noninfective gastroenteritis and colitis |
| K52.9 | Noninfective gastroenteritis and colitis, unspecified |
| R11 | Nausea and vomiting |
| R19.7 | Diarrhea, unspecified |

#### S7.2 GP attendance medical codes for acute kidney injury

| Medcode ID | Code description |
|---|---|
| 2388721000000111 | Acute kidney injury warning stage |
| 2204191000000110 | Acute kidney injury |
| 354406016 | Renal impairment |
| 460091000006111 | Acute renal failure |
| 86957016 | Nephrotic syndrome |
| 2206791000000119 | Acute kidney injury stage 1 |
| 354407013 | Impaired renal function |
| 259688011 | Renal function tests abnormal |
| 2206811000000118 | Acute kidney injury stage 2 |
| 2206831000000114 | Acute kidney injury stage 3 |
| 303943013 | Impaired renal function disorder |
| 459420012 | Deteriorating renal function |
| 619541000006116 | Dialysis for renal failure |
| 354320010 | Nephropathy |
| 303907019 | Glomerulonephritis |
| 303826014 | Nephrotic syndrome with membranous glomerulonephritis |
| 304058013 | Nephritis, nephrosis and nephrotic syndrome NOS |
| 677951000006111 | Nephritis |
| 354417015 | Acute-on-chronic renal failure |
| 303916015 | Acute renal failure NOS |
| 47940012 | Acute interstitial nephritis |
| 303806010 | Nephritis, nephrosis and nephrotic syndrome |
| 678061000006111 | Nephropathy - chronic |
| 303855018 | Nephrotic syndrome NOS |
| 396706011 | Nephrotic syndrome with minimal change glomerulonephritis |
| 303960015 | Impaired renal function disorder NOS |
| 303875010 | Membranous glomerulonephritis |
| 303905010 | Interstitial nephritis |
| 481692010 | Acute renal failure due to rhabdomyolysis |
| 396707019 | Nephritis and nephropathy unspecified |
| 677941000006114 | Nephritis - chronic |
| 303833014 | Nephrotic syndrome, focal and segmental glomerular lesions |
| 460151000006118 | Acute tubular necrosis |
| 14415811000006111 | Acute kidney injury stage 1 |
| 2645796013 | Acute renal failure due to ACE inhibitor |
| 2732241000006113 | Acute renal failure syndrome |
| 886621000006112 | Nephritis NOS |
| 303834015 | Nephrotic syndrome, diffuse membranous glomerulonephritis |
| 461420012 | Dialysis procedure |
| 429641000006113 | Tubulointerstitial nephritis |
| 677461000006115 | Nephrotic syndrome, diffuse mesangial proliferative glomerulonephritis |
| 677561000006119 | Nephrotic syndrome in systemic lupus erythematosus |
| 354413016 | Acute drug-induced renal failure |
| 303846016 | Nephrotic syndrome in diabetes mellitus |
| 303915016 | Other acute renal failure |
| 349998013 | Haemofiltration |
| 303845017 | Nephrotic syndrome in amyloidosis |
| 14415821000006115 | Acute kidney injury stage 2 |
| 14415831000006117 | Acute kidney injury stage 3 |
| 1705781000006113 | Acute renal failure due to obstruction |
| 303839013 | Nephrotic syndrome, diffuse crescentic glomerulonephritis |
| 303832016 | Nephrotic syndrome, minor glomerular abnormality |
| 677471000006110 | Nephrotic syndrome, diffuse mesangiocapillary glomerulonephritis |
| 677621000006112 | Nephrotic syndrome with membranoproliferative glomerulonephritis |
| 481693017 | Acute tubular necrosis |
| 303825013 | Nephrotic syndrome with proliferative glomerulonephritis |
| 303844018 | Nephrotic syndrome in diseases EC |
| 3183501000006111 | Renal failure syndrome |
| 2184271000000115 | Acute renal failure due to non-traumatic rhabdomyolysis |
| 677981000006115 | Nephritis unsp+OS membranoprolif glomerulonephritis lesion |
| 886611000006116 | Nephritis/nephrosis/neph syndr |
| 678081000006118 | Nephropathy induced by other drugs meds and biologl substncs |
| 460081000006113 | Acute renal cortical necrosis |
| 303819014 | Acute diffuse nephritis |

| Medcode ID | Code description |
|---|---|
| 508913018 | Necrotising renal papillitis |
| 1221119015 | ARF - Acute renal failure |
| 405116017 | Acute renal medullary necrosis |
| 1230447010 | Acute idiopathic interstitial nephritis |
| 1847641000006114 | Acute renal failure induced by non-steroidal anti-inflammatory drug |
| 2191301000000116 | Acute renal failure induced by radiographic contrast media |
| 2198081000000118 | Acute renal failure due to traumatic rhabdomyolysis |
| 5986751000006115 | Kidney biopsy sample |
| 303853013 | Nephrotic syndrome associated with another disorder |
| 303951011 | Renal function impairment with growth failure |
| 8040791000006115 | Acute renal failure |
| 460121000006110 | Acute renal failure following labour and delivery |
| 678091000006115 | Nephropathy induced by unspec drug medicament or biol subs |
| 2615221000006111 | Kidney biopsy |
| 677971000006118 | Nephritis unsp+membranoprolif glomerulonephritis lesion NOS |
| 12717101000006112 | Nephritis, nephrosis and nephrotic syndrome |
| 355554015 | Nephropathy NOS in pregnancy without hypertension |
| 303838017 | Nephrotic syndrome, dense deposit disease |
| 2214211000000112 | Acute renal failure induced by toxin |
| 7954321000006110 | Acute renal failure stage 1 |
| 5094041000006117 | Acute renal impairment |
| 303849011 | Nephrotic syndrome in polyarteritis nodosa |
| 303854019 | Nephrotic syndrome with other pathological kidney lesions |
| 405118016 | Acute papillary necrosis |
| 13485211000006111 | Acute kidney injury due to disease caused by SARS-CoV-2 (severe acute respiratory syndrome coronavirus 2) |
| 304053016 | Nephropathy induced by heavy metals |
| 5966771000006111 | Transient acute renal failure |
| 5094071000006113 | Nephrotoxic acute renal failure |
| 305571011 | Acute renal failure following abortive pregnancy |
| 14057041000006114 | Acute tubulointerstitial nephritis |
| 303848015 | Nephrotic syndrome in malaria |
| 677481000006113 | Nephrotic syndrome, diffuse endocapillary proliferative glomerulonephritis |
| 2191151000000117 | Acute renal failure induced by poison |
| 7827241000006112 | Acute kidney injury due to hypovolaemia |
| 7829401000006116 | Acute kidney injury due to sepsis |
| 3136131000006110 | Abnormal renal function |
| 7214431000006112 | Biopsy of kidney using ultrasound guidance |
| 8425521000006110 | Acute kidney injury due to acute tubular necrosis due to sepsis |
| 5093891000006115 | Membranous glomerulonephritis - stage IV |
| 2180741000000113 | Acute renal failure induced by aminoglycoside |
| 2180861000000117 | Acute renal failure induced by cisplatin |
| 2180901000000112 | Acute renal failure induced by cyclosporin A |
| 7100101000006115 | Acute renal failure due to acute cortical necrosis |
| 6927831000006111 | Tubulointerstitial nephritis with uveitis syndrome |
| 4195491000006118 | Hemolytic uremic syndrome |
| 12704041000006118 | Acute renal cortical necrosis |
| 7879771000006116 | Acute kidney injury due to trauma |
| 8425491000006113 | Acute kidney injury due to acute tubular necrosis due to hypovolaemia |
| 13485231000006117 | Acute kidney injury due to disease caused by Severe acute respiratory syndrome coronavirus 2 |
| 2185551000000111 | Acute renal failure induced by animal toxin |
| 2185591000000115 | Acute renal failure induced by plant toxin |
| 2185631000000115 | Acute renal failure induced by heavy metal |
| 2191381000000114 | Acute renal failure induced by solvent |
| 7954341000006115 | Acute renal failure stage 2 |
| 7954361000006116 | Acute renal failure stage 3 |
| 4789251000006116 | Nephropathy associated with vesicoureteral reflux |
| 7841561000006115 | Acute renal insufficiency |
| 5093831000006119 | Membranous glomerulonephritis - stage I |
| 7085691000006115 | Tubulo-interstitial nephritis |

#### S7.3 GP attendance medical codes for gastroenteritis

| Medcode ID | Code description |
|---|---|
| 317494018 | Loose stools |
| 317462011 | [D]Nausea and vomiting NOS |
| 451469011 | [D]Projectile vomiting |
| 317459013 | [D]Emesis |
| 317458017 | Vomiting |
| 317457010 | [D]Nausea |
| 317456018 | Nausea and vomiting |
| 79989019 | Vomiting in newborn |

| Medcode ID | Code description |
|---|---|
| 181648100006111 | Gastroenteritis and colitis of unknown origin |
| 619781000006115 | Diarrhoea - presumed non-infectious |
| 72962011 | Enterocolitis |
| 42550011 | Gastroenteritis |
| 302640019 | Persistent vomiting NOS |
| 302638012 | Cyclical vomiting |
| 302636011 | Persistent vomiting |
| 301423018 | Influenza with gastrointestinal tract involvement |
| 201518100006112 | Norovirus infection |
| 982731000006112 | Diarrhoea/loose stools |
| 854661000006115 | Diarrhoea & vomiting |
| 377011000006119 | Gastroenteritis presumed infectious |
| 287995017 | [X]Other specified intestinal infections |
| 287994018 | Viral and ill-defined gastrointestinal infections |
| 287993012 | Viral enteritis |
| 287990010 | Amoebic infection |
| 287987016 | Amoebic infection |
| 287986013 | [X]Bacterial food-borne intoxication, unspecified |
| 287985012 | [X]Other specified bacterial food-borne intoxications |
| 287984011 | Bacterial enteritis |
| 287983017 | [X]Other specified bacterial intestinal infections |
| 287982010 | [X]Shigellosis, unspecified |
| 287981015 | [X]Other shigellosis |
| 287980019 | [X]Salmonella infection, unspecified |
| 287979017 | [X]Other specified salmonella infections |
| 287977015 | [X]Cholera, unspecified |
| 347485016 | Acanthamoeba keratitis |
| 287944019 | Acanthamoebiasis |
| 123933019 | Winter vomiting disease |
| 123932012 | Epidemic vomiting syndrome |
| 633521000006119 | E. coli infection |
| 501778016 | Escherichia coli infection |
| 286213013 | Intestinal tract infectious disease NOS |
| 286212015 | Other specified infectious diseases of intestinal tract |
| 878891000006118 | Specific GIT infectious dis. |
| 785691000006115 | Ill-defined intestinal infection |
| 878931000006110 | Viral + ill-defined GIT inf. |
| 360012013 | Infantile gastroenteritis |
| 619761000006113 | Infectious diarrhoeal disease |
| 72144015 | Diarrhoea of presumed infectious origin |
| 286202010 | Infectious diarrhoea NOS |
| 179018017 | Viral gastroenteritis |
| 86701000006110 | Traveler's diarrhoea |
| 201288012 | Epidemic diarrhoea |
| 479228018 | Infectious diarrhoea |
| 572061000006117 | Colitis, enteritis and gastroenteritis presumed infect NOS |
| 1494813012 | Gastroenteritis - presumed infectious origin |
| 353405010 | Suspected infectious enteritis |
| 645721000006113 | Enteritis presumed infectious |
| 1495457013 | Colitis - presumed infectious origin |
| 473395016 | Gastric flu |
| 395323013 | Colitis, enteritis and gastroenteritis presumed infectious |
| 286182018 | Gastrointestinal infection |
| 878941000006117 | Gastroenteritis - viral + NOS |
| 21441016 | Infectious gastroenteritis |
| 91741012 | Infectious enteritis |
| 65964017 | Infectious colitis |
| 286180014 | Infectious colitis, enteritis and gastroenteritis |
| 1222445017 | Ill-defined intestinal tract infections |
| 799701000006116 | Infectious disease of digestive tract |
| 451427012 | Infantile viral gastroenteritis |
| 61191000006113 | Viral gastroenteritis |
| 799691000006116 | Gastrointestinal tract infection specified organism NEC |
| 286173012 | Enteritis due to specified virus NOS |
| 117478100000115 | Enteritis due to Norovirus |
| 365356015 | Viral vomiting |
| 286172019 | Enteritis due to rotavirus |
| 365357012 | Viral diarrhoea |
| 1227968013 | Enteritis due to enterovirus |
| 501712011 | Enteritis due to adenovirus |
| 395321010 | Enteritis due to specified virus |
| 286165015 | Unspecified bacterial enteritis |
| 286164016 | Other specified gastrointestinal tract infections NOS |
| 286163010 | Other specified other gastrointestinal infection |

| Medcode ID | Code description |
|---|---|
| 1234534014 | Enteritis due to Yersinia enterocolitica |
| 533191000006119 | Campylobacter intestinal infection |
| 619811000006118 | Diarrhoea due to Campylobacter jejuni |
| 533201000006116 | Enteric campylobacteriosis |
| 619821000006114 | Diarrhoea due to Pseudomonas pyocyanea |
| 395320011 | Pseudomonas gastrointestinal tract infection |
| 619831000006112 | Diarrhoea due to staphylococcal toxin |
| 1773224011 | Diarrhoea due to staphylococcus |
| 395319017 | Staphylococcal gastrointestinal tract infection |
| 286141018 | Bacterial intestinal infectious disease |
| 286140017 | Proteus gastrointestinal tract infection NOS |
| 286139019 | Proteus morganii gastrointestinal tract infection |
| 286137017 | Proteus mirabilis gastrointestinal tract infection |
| 286136014 | Proteus gastrointestinal tract infection |
| 472961000006112 | Gastrointestinal infection caused by Klebsiella aerogenes |
| 286134012 | Arizona paracolon gastrointestinal tract infection |
| 360044012 | Enterohaemorrhagic Escherichia coli infection |
| 360051015 | Enteroinvasive Escherichia coli infection |
| 360040015 | Enterotoxigenic Escherichia coli infection |
| 360043018 | Enteropathogenic Escherichia coli infection |
| 365279016 | Escherichia coli gastrointestinal tract infection |
| 286129013 | Intestinal infection due to other organisms |
| 286128017 | Protozoal intestinal diseases NOS |
| 286127010 | Other specified protozoal intestinal diseases |
| 360073010 | Cryptosporidiosis |
| 572031000006114 | Giardial colitis |
| 802121000006117 | Giardiasis |
| 286119018 | Protozoal intestinal disease |
| 286117016 | Amoebiasis NOS |
| 91483015 | Amoebic nondysenteric colitis |
| 286104019 | Acute amoebic dysentery |
| 1490309018 | Amoebiasis |
| 286102015 | Food poisoning NOS |
| 1222370018 | Foodborne Bacillus cereus intoxication |
| 286100011 | Other specified bacterial food poisoning |
| 1234561014 | Vibrio parahaemolyticus food poisoning |
| 286099015 | Other Clostridia causing food poisoning |
| 569601000006113 | Clostridium perfringens food poisoning |
| 1234964014 | Staphylococcal food poisoning |
| 286096010 | Other bacterial food poisoning |
| 286095014 | Shigellosis NOS |
| 286094013 | Other specified shigella infection |
| 878911000006116 | Bacillary dysentry |
| 501911000006117 | Bacillary dysentery Shigella sonnei |
| 395316012 | Shigella sonnei |
| 286089013 | Shigella boydii (group C) |
| 286088017 | Shigella flexneri |
| 988691000006119 | Bacillary dysentry |
| 409836012 | Bacillary dysentery |
| 395315011 | Shigella dysenteriae |
| 353844012 | Vomiting co-occurrent and due to infectious disease |
| 60387019 | Shigellosis |
| 286084015 | Salmonella infection |
| 286083014 | Other specified salmonella infection |
| 286078018 | Local salmonella infection unspecified |
| 494811016 | Localised Salmonella infection |
| 443811013 | Salmonella food poisoning |
| 443815016 | Salmonellosis |
| 443812018 | Salmonella gastroenteritis |
| 124991010 | Food poisoning |
| 286071012 | Other salmonella infections |
| 109787018 | Bacterial food poisoning |
| 286066017 | Typhoid and paratyphoid fevers |
| 286065018 | Cholera NOS |
| 126156018 | Vibrio cholerae |
| 551671000006114 | Cholera - Vibrio cholerae El Tor |
| 551661000006119 | Cholera - Vibrio cholerae |
| 105789012 | Cholera |
| 222831000000115 | Intestinal infectious disease |
| 763991000006113 | Food poisoning notification administration |
| 264402018 | Notification of food poisoning |
| 261410016 | Stool culture positive |
| 261408018 | Stool culture cryptosporidium positive |
| 261251016 | Sample salmonella cultured |

| Medcode ID | Code description |
|---|---|
| 1539981000006116 | Norovirus (small round-structured viruses) nucleic acid detn |
| 1923531000006118 | Rotavirus RNA (ribonucleic acid) detection assay |
| 2335181000000114 | Rotavirus nucleic acid detection assay |
| 313311000000111 | Rotavirus nucleic acid detection |
| 1923481000006116 | Norovirus RNA (ribonucleic acid) detection assay |
| 2333361000000116 | Norovirus nucleic acid detection assay |
| 313191000000117 | Norovirus nucleic acid detection |
| 1488689010 | Suspected food poisoning |
| 372283012 | Diarrhoea and vomiting |
| 619771000006118 | Diarrhoea and vomiting, symptom |
| 397928013 | Diarrhoea symptom |
| 103081000006118 | Toddler diarrhoea |
| 1786047017 | Loose stool |
| 103578017 | Diarrhoea |
| 619741000006114 | Diarrhoea |
| 397927015 | Diarrhoea symptoms |
| 252622013 | Vomiting NOS |
| 2238541000000114 | Frequency of vomiting |
| 252620017 | Bilious vomiting |
| 372248013 | Vomiting symptom |
| 15155012 | Projectile vomiting |
| 100841000006111 | Throwing up |
| 2647992016 | Emesis |
| 2643931013 | Vomiting |
| 407086011 | C/O - vomiting |
| 63631000006119 | Finding of vomiting |
| 5730511000006110 | Diarrhoea and vomiting after gastrointestinal tract surgery |
| 5089301000006119 | Diarrhoea due to ingestion of unabsorbable substances |
| 13012291000006117 | Gastroenteritis caused by SARS-CoV-2 (severe acute respiratory syndrome coronavirus 2) |
| 12990711000006115 | Gastroenteritis caused by 2019-nCoV (novel coronavirus) |
| 3512083018 | Diarrhoea co-occurrent and due to carcinoid syndrome |
| 6746021000006114 | Norovirus |
| 7303631000006118 | Norovirus infection |

#### S8. Supplement references

1. Harris JP, Iturriza-Gomara M, O'Brien SJ. Re-assessing the total burden of norovirus circulating in the United Kingdom population. Vaccine. 2017 Feb;35(6):853–5.

2. Simmons K, Gambhir M, Leon J, Lopman B. Duration of Immunity to Norovirus Gastroenteritis. Emerg Infect Dis. 2013 Aug;19(8):1260–7.

3. Steele MK, Remais JV, Gambhir M, Glasser JW, Handel A, Parashar UD, et al. Targeting pediatric versus elderly populations for norovirus vaccines: a model-based analysis of mass vaccination options. Epidemics. 2016 Dec;17:42–9.

4. Sukhrie FHA, Teunis P, Vennema H, Copra C, Thijs Beersma MFC, Bogerman J, et al. Nosocomial Transmission of Norovirus Is Mainly Caused by Symptomatic Cases. Clin Infect Dis. 2012 Apr 1;54(7):931–7.

5. Gaythorpe KAM, Trotter CL, Conlan AJK. Modelling norovirus transmission and vaccination. Vaccine. 2018 Sep;36(37):5565–71.

6. Glass RI, Parashar UD, Estes MK. Norovirus Gastroenteritis. N Engl J Med. 2009 Oct 29;361(18):1776–85.

7. Atmar RL, Opekun AR, Gilger MA, Estes MK, Crawford SE, Neill FH, et al. Norwalk Virus Shedding after Experimental Human Infection. Emerg Infect Dis. 2008 Oct;14(10):1553–7.

8. Wu Q song, Xuan Z liang, Liu J yi, Zhao X tao, Chen Y fang, Wang C xi, et al. Norovirus shedding among symptomatic and asymptomatic employees in outbreak settings in Shanghai, China. BMC Infect Dis. 2019 Dec;19(1):592.

9. Office for National Statistics. Vital statistics in the UK: births, deaths and marriages [Internet]. Available from: https://www.ons.gov.uk/peoplepopulationandcommunity/populationandmigration/populationestimates/datasets/vitalstatisticspopulationandhealthreferencetables

10. Mossong J, Hens N, Jit M, Beutels P, Auranen K, Mikolajczyk R, et al. Social Contacts and Mixing Patterns Relevant to the Spread of Infectious Diseases. Riley S, editor. PLoS Med. 2008 Mar 25;5(3):e74.

11. Parrino TA, Schreiber DS, Trier JS, Kapikian AZ, Blacklow NR. Clinical Immunity in Acute Gastroenteritis Caused by Norwalk Agent. N Engl J Med. 1977 Jul 14;297(2):86–9.

12. O'Reilly KM, Sandman F, Allen D, Jarvis CI, Gimma A, Douglas A, et al. Predicted norovirus resurgence in 2021–2022 due to the relaxation of nonpharmaceutical interventions associated with COVID-19 restrictions in England: a mathematical modeling study. BMC Med. 2021 Dec;19(1):299.

13. Tam CC, Rodrigues LC, Viviani L, Dodds JP, Evans MR, Hunter PR, et al. Longitudinal study of infectious intestinal disease in the UK (IID2 study): incidence in the community and presenting to general practice. Gut. 2012 Jan;61(1):69–77.

14. Verstraeten T, Cattaert T, Harris J, Lopman B, Tam CC, Ferreira G. Estimating the Burden of Medically Attended Norovirus Gastroenteritis: Modeling Linked Primary Care and Hospitalization Datasets. J Infect Dis. 2017 Nov 15;216(8):957–65.

15. Sandmann FG, Shallcross L, Adams N, Allen DJ, Coen PG, Jeanes A, et al. Estimating the Hospital Burden of Norovirus-Associated Gastroenteritis in England and Its Opportunity Costs for Nonadmitted Patients. Clin Infect Dis. 2018 Aug 16;67(5):693–700.

16. Public Health England. PHE National norovirus and rotavirus Report 09 April 2020 – Week 15 report (data to week 13) [Internet]. Available from: https://www.gov.uk/government/statistics/norovirus-and-rotavirus-summary-of-surveillance-2019-to-2020

17. Ondrikova N, Clough HE, Cunliffe NA, Iturriza-Gomara M, Vivancos R, Harris JP. Understanding norovirus reporting patterns in England: a mixed model approach. BMC Public Health. 2021 Dec;21(1):1245.

18. Clinical Practice Research Datalink [Internet]. 2021. Available from: https://cprd.com/Data

19. Wolf A, Dedman D, Campbell J, Booth H, Lunn D, Chapman J, et al. Data resource profile: Clinical Practice Research Datalink (CPRD) Aurum. Int J Epidemiol. 2019 Dec 1;48(6):1740–1740g.

20. Bhaskaran K, Gasparrini A, Hajat S, Smeeth L, Armstrong B. Time series regression studies in environmental epidemiology. Int J Epidemiol. 2013 Aug;42(4):1187–95.

21. NHS England and NHS Improvement. National Cost Collection for the NHS [Internet]. Available from: https://www.england.nhs.uk/costing-in-the-nhs/national-cost-collection/

22. Department of Health and Social Care. NHS reference costs [Internet]. Available from: https://www.gov.uk/government/collections/nhs-reference-costs
